# A machine learning approach identifies unresolving secondary pneumonia as a contributor to mortality in patients with severe pneumonia, including COVID-19

**DOI:** 10.1101/2022.09.23.22280118

**Authors:** Catherine A. Gao, Nikolay S. Markov, Thomas Stoeger, Anna Pawlowski, Mengjia Kang, Prasanth Nannapaneni, Rogan A. Grant, Chiagozie Pickens, James M. Walter, Jacqueline M. Kruser, Luke Rasmussen, Dan Schneider, Justin Starren, Helen K. Donnelly, Alvaro Donayre, Yuan Luo, GR Scott Budinger, Richard G. Wunderink, Alexander V. Misharin, Benjamin D. Singer, The NU SCRIPT Study Investigators

**Affiliations:** Division of Pulmonary and Critical Care Medicine, Department of Medicine, Northwestern University Feinberg School of Medicine, Chicago, IL; Department of Chemical and Biological Engineering, Northwestern University, McCormick School of Engineering, Evanston, IL; Northwestern Medicine Enterprise Data Warehouse, Northwestern University Feinberg School of Medicine, Chicago, IL; Division of Allergy, Pulmonary and Critical Care, Department of Medicine, University of Wisconsin School of Medicine and Public Health, Madison, Wisconsin; Division of Health and Biomedical Informatics, Department of Preventive Medicine, Northwestern University Feinberg School of Medicine, Chicago, IL; Simpson Querrey Lung Institute for Translational Science at Northwestern University (SQ^LIFTS^NU)

## Abstract

**Background:** Patients with severe SARS-CoV-2 pneumonia experience longer durations of critical illness yet similar mortality rates compared to patients with severe pneumonia secondary to other etiologies. As secondary bacterial infection is common in SARS-CoV-2 pneumonia, we hypothesized that unresolving ventilator-associated pneumonia (VAP) drives the apparent disconnect between length-of-stay and mortality rate among these patients.

**Methods:** We analyzed VAP in a prospective single-center observational study of 585 mechanically ventilated patients with suspected pneumonia, including 190 patients with severe SARS-CoV-2 pneumonia. We developed *CarpeDiem*, a novel machine learning approach based on the practice of daily ICU team rounds to identify clinical states for each of the 12,495 ICU patient-days in the cohort. We used the *CarpeDiem* approach to evaluate the effect of VAP and its resolution on clinical trajectories.

**Findings:** Patients underwent a median [IQR] of 4 [2,7] transitions between 14 clinical states during their ICU stays. Clinical states were associated with differential hospital mortality. The long length-of-stay among patients with severe SARS-CoV-2 pneumonia was associated with prolonged stays in clinical states defined by severe respiratory failure and with a lower frequency of transitions between clinical states. In all patients, including those with COVID-19, unresolving VAP episodes were associated with transitions to unfavorable states and hospital mortality.

**Interpretation:** *CarpeDiem* offers a machine learning approach to examine the effect of VAP on clinical outcomes. Our findings suggest an underappreciated contribution of unresolving secondary bacterial pneumonia to outcomes in mechanically ventilated patients with pneumonia, including due to SARS-CoV-2.

Graphical abstract
**Disentangling the contributions of ICU complications and interventions to ICU outcomes**. (**A**) Traditional approaches evaluate the ICU stay as a black box with severity of illness measured on presentation and dichotomized survival at an arbitrary time point (e.g., day 28) or on ICU or hospital discharge. Hence, the effect of intercurrent complications and interventions cannot be easily measured, a problem that is compounded when ICU stays are long or significantly differ between groups. (**B**) Defining the ICU course by clinical features during each day in the ICU permits the association of a complication or intervention with transitions toward clinical states associated with favorable or unfavorable outcomes.

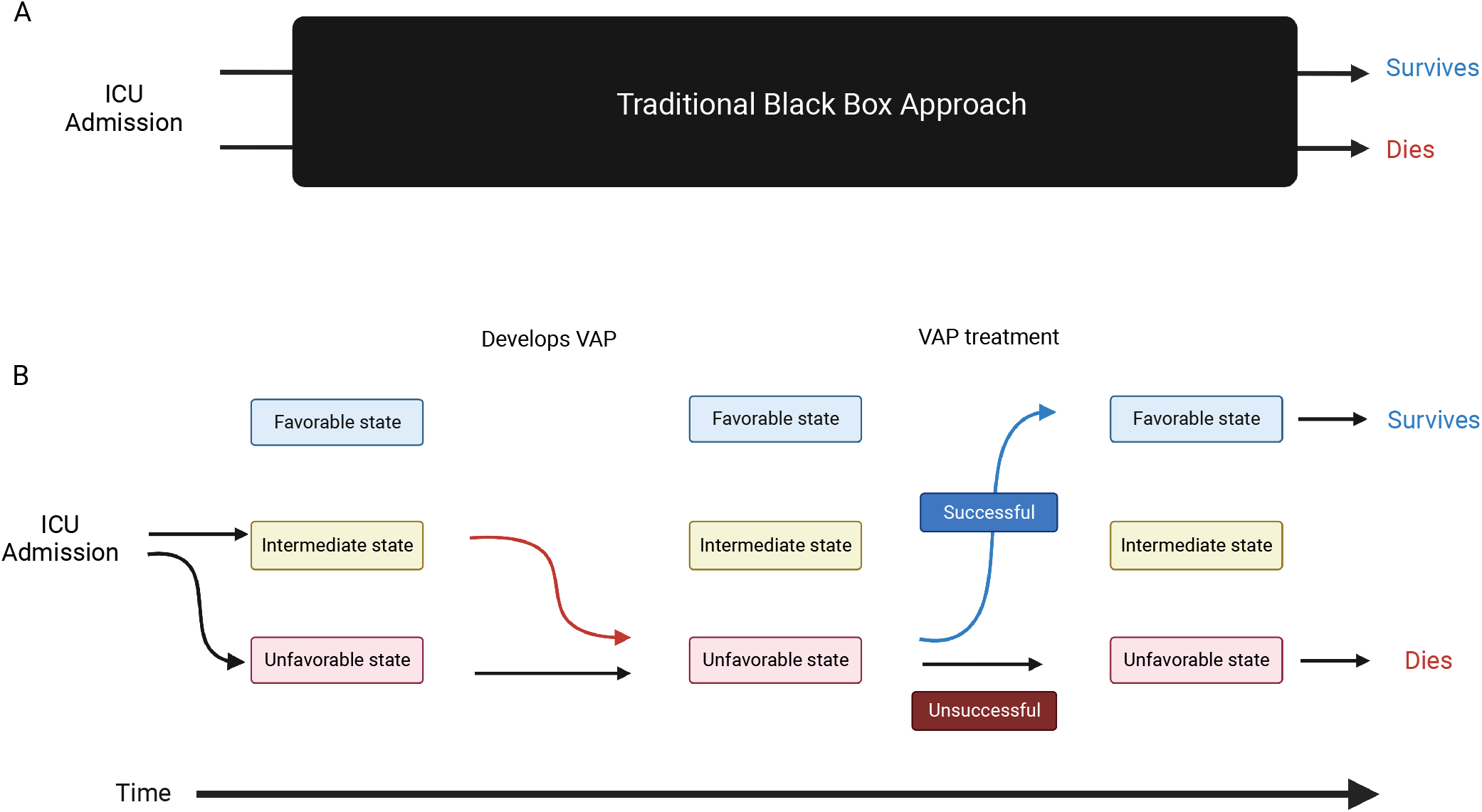

## Introduction

In a review of autopsy samples stored from the 1918 influenza A pandemic, Fauci and colleagues suggested an unexpectedly important contribution of secondary bacterial infection to mortality after severe viral pneumonia.^1^ We observed high rates of ventilator-associated pneumonia (VAP) in patients with severe acute respiratory syndrome coronavirus-2 (SARS-CoV-2) pneumonia requiring mechanical ventilation, suggesting bacterial superinfections may also contribute to mortality in these patients.^2^ In parallel, we and others observed that duration of ICU stay and mechanical ventilation is more than twice as long in patients with SARS-CoV-2 pneumonia compared to patients with respiratory failure complicating pneumonia caused by other pathogens and patients with other causes of the acute respiratory distress syndrome (ARDS).^3–8^ Despite longer lengths of stay (LOS), mortality is similar in patients with severe SARS-CoV-2 pneumonia and those with pneumonia secondary to other pathogens.^4–6,8^ These findings support a paradigm in which a relatively low mortality directly attributable to primary SARS-CoV-2 infection is offset by an increased risk of death attributable to unresolving secondary bacterial pneumonia.^9^

Testing the hypothesis that unresolving secondary bacterial pneumonia explains the apparent disconnect between LOS and mortality in patients with severe SARS-CoV-2 pneumonia poses two challenges. First, traditional methods to compare ICU outcomes standardize severity of illness on ICU admission and treat the entirety of the ensuing ICU stay as a singular event, neglecting the effect and timing of ICU complications that develop or therapies that are administered during the ICU stay on outcomes. This approach fails to capture ICU complications, such as VAP, that by definition are not present on admission but likely alter the trajectory of the patient toward unfavorable outcomes, particularly when ICU LOS is long. Indeed, few ICU studies have attempted to examine the effect of late ICU interventions and complications on patient outcomes.^10–12^ Second, VAP episodes may contribute to prolonged LOS while not worsening outcomes if they resolve. Nevertheless, most studies use insensitive methods to diagnose secondary bacterial pneumonia and measure response to therapy.^13^

To disentangle the effect of intercurrent ICU events such as secondary bacterial pneumonia on outcomes, we analyzed the contribution of VAP to mortality in 585 patients with severe pneumonia and respiratory failure, including 190 patients with severe SARS-CoV-2 pneumonia, enrolled in the Successful Clinical Response in Pneumonia Therapy (SCRIPT) study. All patients underwent bronchoalveolar lavage (BAL) sampling paired with comprehensive microbiological diagnostics as part of routine clinical care whenever pneumonia was suspected. Clinicians used the results of these studies to guide antimicrobial therapy.^2^ To understand the contribution of VAP to outcomes over the course of the ICU stay, we developed a machine learning approach, *CarpeDiem*, which revealed that unresolving episodes of VAP are associated with transitions to progressively unfavorable clinical states and increased mortality. Our findings suggest an unexpected contribution of unresolving VAP to mortality in patients with severe pneumonia, including COVID-19, and provide a machine learning method to analyze contributions of ICU complications and treatments to patient outcomes.

## Methods

### Study setting

Patients were enrolled in the Successful Clinical Response in Pneumonia Therapy (SCRIPT) Systems Biology Center, a single-site, prospective cohort study of patients hospitalized in the ICUs of Northwestern Memorial Hospital (NMH) with suspected severe pneumonia (defined as requiring mechanical ventilation), all of whom underwent at least one BAL procedure. This study was approved by the Northwestern University Institutional Review Board with study ID STU00204868.

### Study procedures

ICU physicians at NMH routinely obtain bronchoscopic or non-bronchoscopic BAL samples from mechanically ventilated patients with suspected pneumonia.^14^ NMH clinical laboratories performed quantitative bacterial culture and antimicrobial sensitivity testing. Many of the samples were also analyzed by multiplex PCR (BioFire® FilmArray® Pneumonia (PN) Panel) with results provided to clinical teams within 3 hours. We previously reported that our physicians initiate guideline-recommended antimicrobial therapy when pneumonia is suspected and use data obtained from analysis of BAL fluid to appropriately narrow or discontinue empirical guideline-recommended antimicrobial therapy.^2^ BAL fluid cell count and differential^8,15^ and serum levels of procalcitonin^16^ were used to assess the pretest probability of pneumonia secondary to bacterial pathogens. BAL fluid amylase was measured to estimate the risk of aspiration.^17^ Physicians were encouraged to order urine testing for *S. pneumoniae* and *L. pneumophila* antigens. BAL fluid acid-fast bacilli and fungal culture and aspergillus galactomannan levels were measured when clinically appropriate.

### Data extraction and analysis

Demographics, clinical data, and outcomes were extracted from the electronic health record (EHR) via the Northwestern Medicine Enterprise Data Warehouse.^18^ For the *CarpeDiem* machine learning approach, we used the following unsupervised strategy, referred to as “Similarity.” First, we manually selected 44 clinical features that we considered representative of those that physicians would consider during daily ICU rounds, including the components of the Sequential Organ Failure Assessment (SOFA) score. Second, we masked some values that we considered out of range (see code for full details). Third, we summarized all values per patient per day, using average or worst value, depending on the feature, and marking absent values as NA. Fourth, we performed pairwise Pearson correlations among all per-day features and considered features with a Pearson correlation above 0.7 to be highly related. Fifth, we normalized each feature with percentile normalization and set tied values to the average percentiles. Sixth, we combined highly related features to a single feature by taking the mean percentile across the related features. Seventh, we created a similarity matrix between all pairs of patient-days by computing the Pearson correlation among features that had values present for both patient-days. Finally, we computed Euclidean distances on the similarity matrix and performed hierarchical clustering on individual patient-days using Ward’s approach.^19^ Additionally, we considered two other strategies: “Ranked-Euclidean” and “Normalized-Euclidean.” During the sixth step, “Ranked-Euclidean” reweights highly related features by dividing their values by the square root of the number of related features, then skips the seventh step, and during the eighth step applies an NA-robust implementation of Euclidean distances as described by Dixon,^20^ available in the *scikit-learn* package.^21^ “Normalized-Euclidean” follows the procedure of “Ranked-Euclidean,” but at the fifth step replaces a general percentile normalization by winsorizing, followed by log_2_ or percentile normalization, depending on specific data input (see code for full details). We validated the *CarpeDiem* approach externally in a suspected pneumonia cohort derived from the MIMIC-IV database,^22^ using code from the MIMIC Code Repository.^23^ We used XGBoost^24^ to model outcomes based on clinical features from early in admission, similar to most clinical prediction models. See Supplemental Materials for extended methods.

### Definition of pneumonia episodes

A panel of six critical care physicians used a prospectively generated, standardized score sheet (Supplemental File 1) to manually review each patient’s EHR, including clinical notes, to identify and categorize pneumonia episodes and adjudicate whether these episodes were successfully treated (resolved). Each case was reviewed independently by two physicians with discrepant adjudications settled by a third. If the third review remained discrepant, the episode was discussed at a meeting of the panelists to determine a consensus decision. Pneumonia episodes were captured up to 99 days following the enrollment BAL procedure and categorized as non-pneumonia controls, other pneumonia (bacterial), other viral pneumonia, or COVID-19. By definition, VAP occurred after at least 48 hours of intubation;^16^ only bacterial VAPs were included in the analysis. VAP duration was defined as the time interval between the diagnostic BAL procedure and clinical cure, discontinuation of antibiotics, or death, whichever was the shortest. Outcomes for VAP episodes were adjudicated at day 7-8, day 10, and day 14 following the diagnostic BAL procedure. Cure was defined as the ability to survive beyond the duration of antibiotic treatment, the ability to remain off of antibiotics for 48 hours without recurrence or superinfection pneumonia, disappearance of the causative pathogen from BAL fluid or the absence of subsequent samples, absence of bacterial complications (e.g., empyema, lung abscess, endocarditis), and improvement in the clinical manifestations of pneumonia; successful extubation or ventilator liberation was considered as a cure. Unsuccessful treatment was defined as at least partial persistence of clinical manifestations or recurrence, persistence, or superinfection (see detailed definitions in Supplemental Materials). Outcomes were classified as indeterminate when partial persistence of clinical manifestations of infection, such as persistence of BAL neutrophilia without positive BAL cultures or PCR, or if antibiotic treatment of a non-pulmonary infection precluded clear assessment of response to pneumonia.

### Statistical analysis

Numerical values were compared using Mann-Whitney U tests with false-discovery rate (FDR) correction with the Benjamini-Hochberg procedure. Categorical values were compared using Fisher’s Exact tests with FDR correction. A p-value or q-value < 0.05 was our threshold for statistical significance.

### Code and data availability

Programming was done in Python (version 3.9). A detailed description of all data extraction and computational procedures, including code, are available at https://github.com/NUSCRIPT/carpediem and in Supplemental Materials. A de-identified version of all SCRIPT cohort data used in this manuscript will be available on PhysioNet.^25^ An interactive data browser illustrating the features of *CarpeDiem* is available on our website, https://nupulmonary.org/carpediem, and will be available through PhysioNet.

## Results

### Demographics

Of 601 patients enrolled in SCRIPT between June 2018 and March 2022, 585 had an adjudicated pneumonia category at the time of analysis: 190 had COVID-19, 50 had pneumonia secondary to other respiratory viruses, 252 had other pneumonia (bacterial), and 93 were initially suspected of having pneumonia yet subsequently adjudicated as having respiratory failure unrelated to pneumonia (non-pneumonia controls). Except for BMI, demographics such as age and gender were similar between groups (Figure 1A-C and Supplemental Table 1). Severity of illness as measured by the Acute Physiology Score (APS) from APACHE IV^26^ and the SOFA score^27,28^ in the first two days of admission did not differ between groups (Figure 1D-E). Despite similar severity of illness on admission, durations of intubation and ICU stay were more than twice as long among patients with SARS-CoV-2 pneumonia compared with any other group, reflected by an increased frequency of tracheostomy (Figure 1F-H and Supplemental Figure 1). Hospital mortality did not differ between groups (Figure 1I).

**Figure 1.**
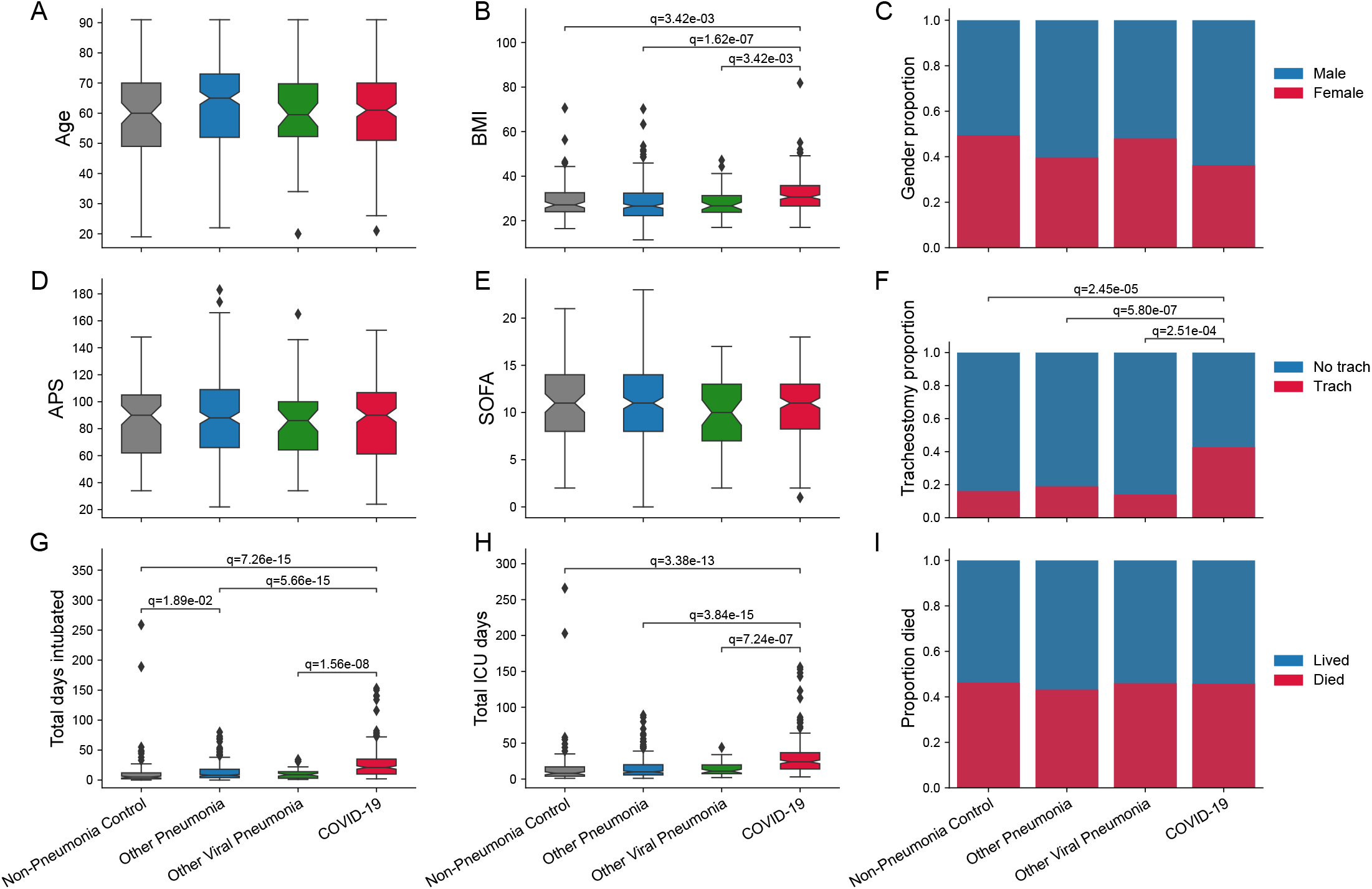
Demographics and outcomes of the cohort grouped by pneumonia category. Distribution of (**A**) patient age, (**B**) BMI,* (**C**) gender, (**D**) APS, (**E**) SOFA score, (**F**) tracheostomy placement, (**G**) duration of intubation**, (**H**) length of ICU stay**, and (**I**) mortality***. * One patient did not have BMI data available. ** Total days intubated (G) and total ICU days (H) include only days at our hospital and do not capture intubation duration or ICU LOS at a transferring hospital. *** Lived includes dispositions of discharge to home, acute inpatient rehabilitation, a long-term acute care hospital (LTACH), or a skilled nursing facility (see Supplemental Table 2). Died includes patients who died, underwent lung transplantation for refractory respiratory failure, or who were transferred to home or inpatient hospice. BMI = body mass index, APS = Acute Physiology Score from APACHE IV (score calculated from worst value within the first two ICU days), SOFA = Sequential Organ Failure Assessment (score calculated from worst value within the first two ICU days).

### CarpeDiem

*a machine learning approach to time-series data in the ICU*. To address the challenge of comparing intercurrent ICU events between groups with different ICU LOS, we developed a machine learning approach, *CarpeDiem*, to discretize each patient day in the ICU. For all 12,495 ICU patient-days in the cohort, we extracted clinical data from the EHR describing 44 parameters, including flags for organ failures requiring mechanical support (e.g., mechanical ventilation, renal replacement therapy, and extracorporeal membrane oxygenation [ECMO]), continuously recorded clinical parameters (e.g., vital signs and doses of norepinephrine), and commonly measured laboratory values (Supplemental Figure 2). Variables used to calculate the SOFA score are a subset of these parameters. Correlation analysis identified expected associations between mathematically or physiologically coupled variables (e.g., plateau pressure, positive end-expiratory pressure [PEEP], and lung compliance; PaCO_2_ and bicarbonate) and revealed clinically recognizable correlated features (e.g., ECMO, D-dimer, and lactate dehydrogenase [LDH]) (Supplemental Figure 2). After reducing the weight of these highly correlated features, we performed clustering using several methods, all of which yielded similar results (Supplemental Figure 3A-C). We designed a clustering strategy based on the similarity between patient-days (see details in Supplemental Methods) and selected the number of clusters by choosing a near-maximal difference in mortality between pairwise comparisons of clusters (Supplemental Figure 4) while clustering only on parameters that were determined to be meaningful by four ICU physicians (CAG, GRSB, RGW, BDS). We visualized the resulting 14 clusters using heatmap and UMAP plots (Figure 2A-C, Supplemental Figure 5).

**Figure 2.**
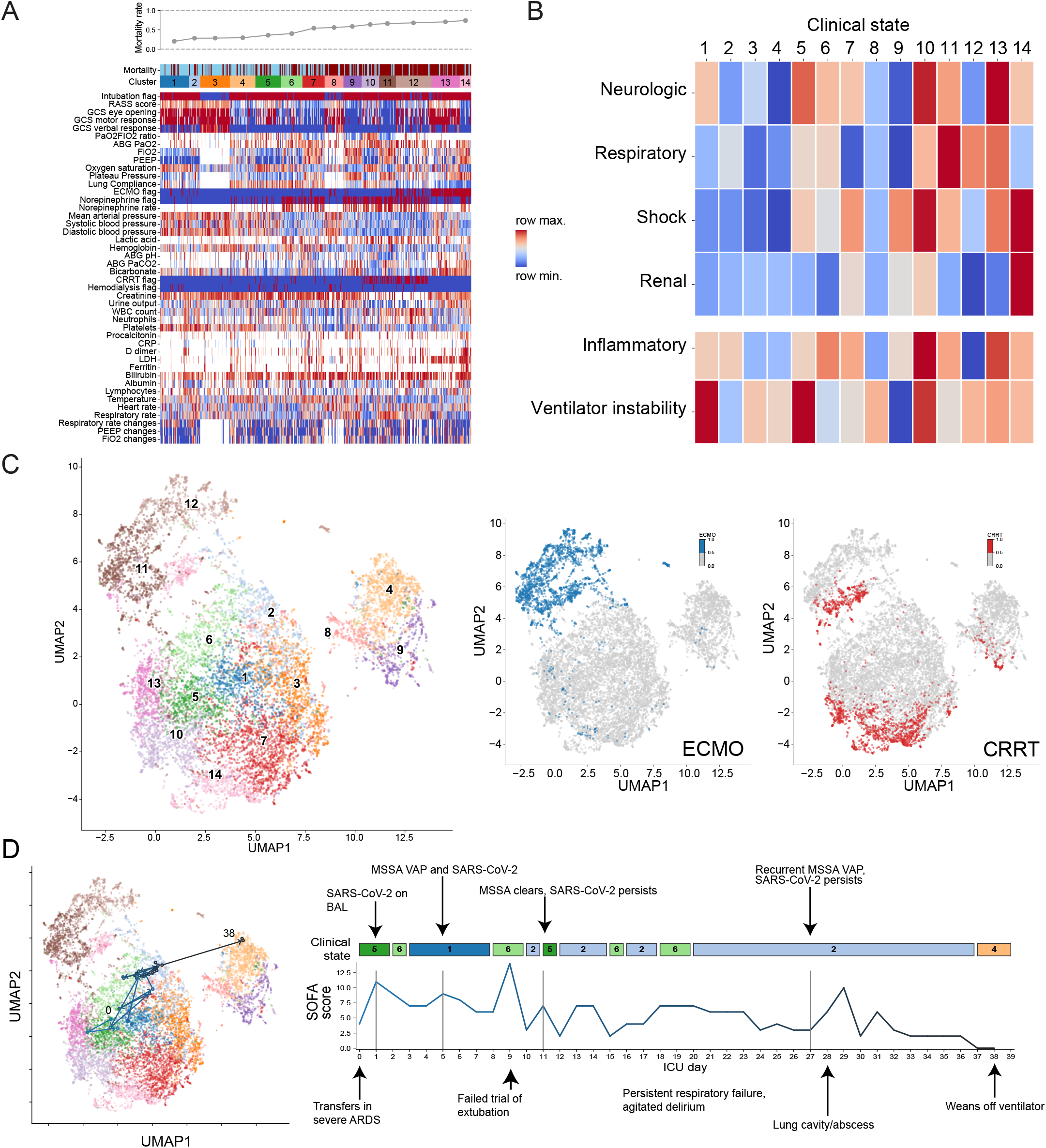
*CarpeDiem* groups patient-days into clusters representing clinical states associated with differential hospital mortality. (**A**) Heatmap of 44 clinical parameters with columns (representing 12,495 ICU patient-days from 585 patients) grouped into *CarpeDiem*-defined clusters (clinical states) ordered from lowest to highest mortality. Rows are sorted into physiologically related groups. The top row signifies hospital mortality of the patient shown in the column (blue = lived, red = died). The hospital mortality rate associated with each cluster is shown above the heatmap. (**B**) Heatmap of the composite signal from each cluster and physiological group with ordering same as (A). (**C**) UMAP with colors and numbers representing *CarpeDiem*-defined clinical states. Feature plots on the right show patient-days spent receiving ECMO (left) or CRRT (right). (**D**) Example trajectory of a patient who was liberated from mechanical ventilation and was discharged home. A UMAP on the left demonstrating clinical transitions is labeled with the starting clinical state (day 0) and the day of discharge from the ICU (day 38). On the right, microbiological and clinical events overlaid on clinical state transitions are shown. Vertical lines indicate timepoints of BAL sampling. The beginning of the ICU course is outlined in light blue, whereas the end is dark blue, on both the timeline and the UMAP. More example trajectories are available in an interactive web app available at https://nupulmonary.org/carpediem. HFNC = high-flow nasal cannula, ECMO = extracorporeal membrane oxygenation, CRRT = continuous renal replacement therapy, MSSA = methicillin-sensitive *Staphylococcus aureus*.

To understand the clinical meaning of the clusters identified by our *CarpeDiem* approach, we arranged the parameters into six physiological groups (neurologic, respiratory, shock, renal, inflammatory, and ventilator instability) and sorted the clusters in order of increasing mortality. The resulting heatmaps revealed an association between patient-days characterized by multiple organ failure and mortality (Figure 2A-B), findings consistent with previously published scoring systems.^28–30^ Moreover, the clusters recapitulated patterns consistent with recognizable clinical states. For example, cluster 11 represents patient-days with very severe respiratory failure (mostly days spent receiving ECMO support), moderately high levels of sedation, an intermediate level of shock without substantial renal failure, and with relatively stable ventilator settings. Hence, the machine learning-defined clusters represent states recognizable to ICU practitioners, and we refer to them as clinical states hereafter. An illustration of time series data and transitions between clinical states over a selected patient’s ICU course is provided in Figure 2D.

We compared mortality for each clinical state identified by *CarpeDiem* on the first and last day in the ICU. On the first day of the ICU stay, only three of the clinical states were significantly associated with outcome (Figure 3A). In contrast, the same analysis for the last ICU day for each patient revealed 10 significant associations between clinical state and outcome (Figure 3B). Nearly all patients (99.9%) underwent transitions between clinical states over the course of their ICU stay (median [IQR] of 4 [2,7] transitions per patient). We defined transitions as favorable if the mortality associated with the destination clinical state was lower than the originating state and *vice versa*.

### *Validation of the* CarpeDiem *approach in the MIMIC-IV dataset*

We next determined whether the *CarpeDiem* approach could be meaningfully applied to an external dataset. Within the MIMIC-IV database of ICU patients, we identified the subset of 1,284 ICU stays similar to those in our cohort. Criteria included admission to and discharge from a medical ICU, respiratory failure requiring mechanical ventilation, and pneumonia as defined by ICD-9 codes. The *CarpeDiem* approach applied to 15,642 ICU patient-days using 27 clinical parameters identified 12 clusters (Supplemental Figure 6A-C). Similar to our observations in the SCRIPT cohort, clinical states in MIMIC-IV were clinically recognizable with increasing organ failure associated with mortality (Supplemental Figure 6D), supporting the generalizability of the *CarpeDiem* approach.

CarpeDiem *reveals that the long LOS in patients with COVID-19 is associated with prolonged stays in clinical states characterized by severe respiratory failure*. We reasoned that *CarpeDiem* could provide insight into the observation that patients with SARS-CoV-2 pneumonia have longer ICU LOS relative to patients with pneumonia secondary to other pathogens despite similar hospital mortality. We posited that this observation could result from either (1) longer stays in a given clinical state with similar numbers of transitions between states or (2) similar durations of stay in any given clinical state with a balanced increase in the number of transitions between favorable and unfavorable states. Although the absolute number of transitions between clinical states was higher among patients with SARS-CoV-2 pneumonia when compared with all other patient groups in the cohort (Figure 4A), the frequency of transitions was significantly lower (Figure 4B). The longer LOS experienced by patients with severe SARS-CoV-2 pneumonia resulted from prolonged stays in seven clinical states (Figure 4C). Time spent in clinical state 11, characterized by severe hypoxemic respiratory failure, accounted for 29.9% of the difference in ICU LOS experienced by patients with COVID-19. While the number of transitions toward favorable outcomes was similar in patients with SARS-CoV-2 pneumonia and other patients in the cohort; the number of favorable transitions was nominally lower in patients with SARS-CoV-2 pneumonia (Figure 5, Supplemental Figure 7).

**Figure 3.**
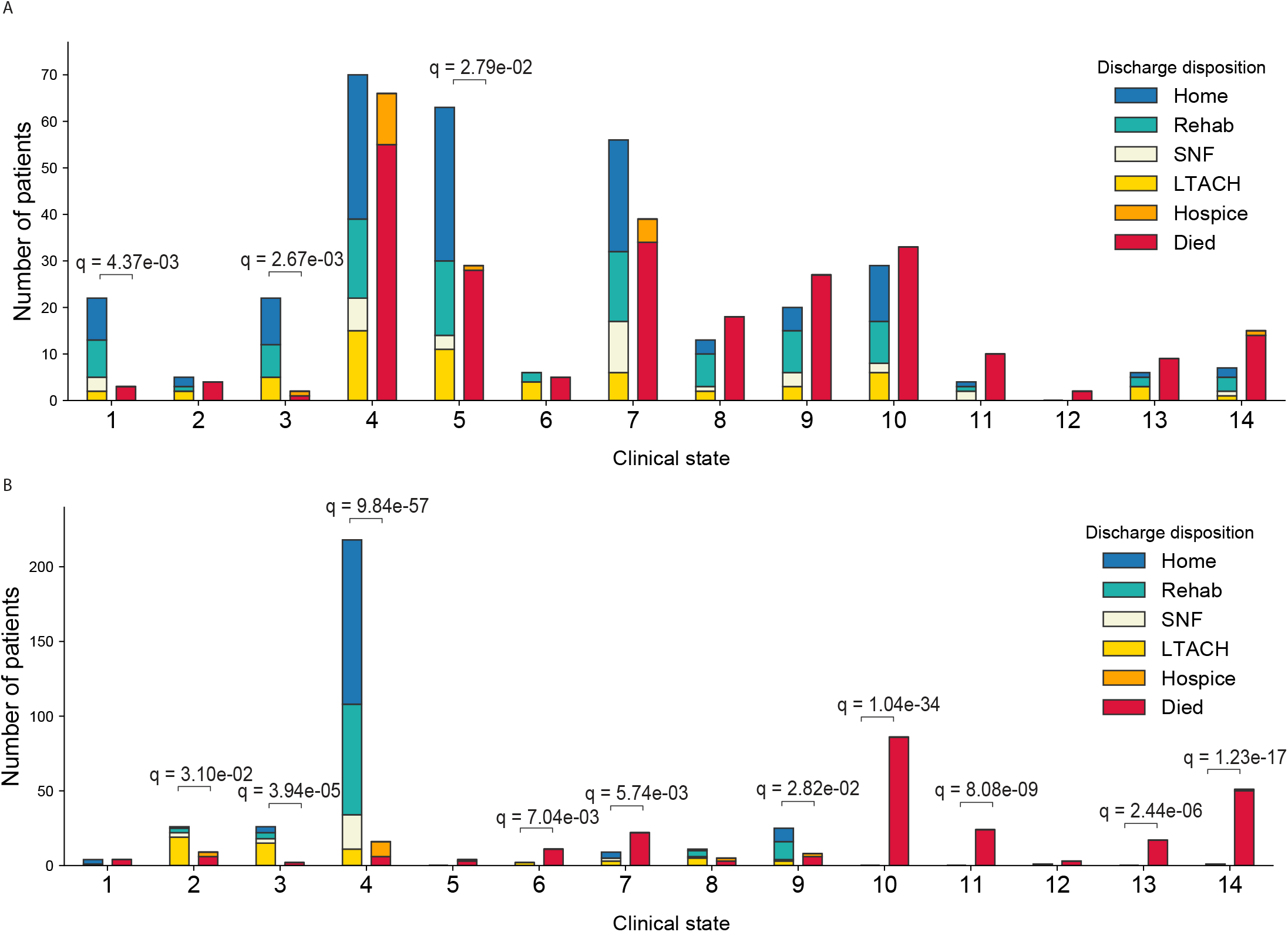
Clinical states are associated with outcome. (**A**) Association between the first clinical state occupied by each patient on their first ICU day and their discharge disposition. (**B**) Association between the last clinical state occupied by each patient on their last ICU day and their discharge disposition. Outcomes are displayed in two columns: the first column aggregates favorable discharge dispositions (Home, Rehab, SNF, LTACH), the second column aggregates unfavorable discharge dispositions (Hospice, Died). SNF = Skilled Nursing Facility, LTACH = long-term acute care hospital.

**Figure 4.**
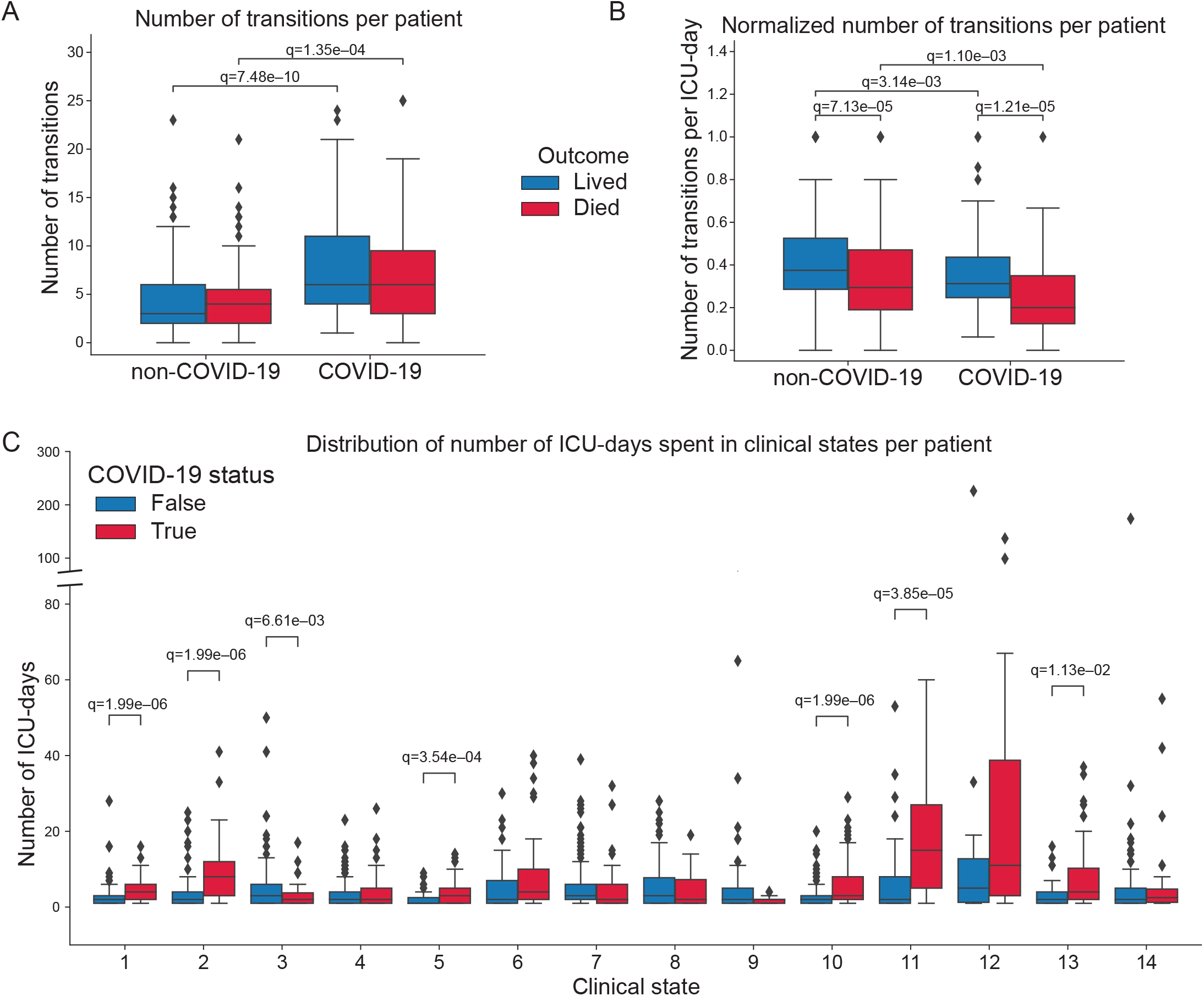
The long length of stay among patients with COVID-19 is driven by longer durations of time spent in clinical states characterized by severe respiratory failure with a lower frequency of transitions. (**A**) Distribution of transitions per patient. (**B**) Distribution of transitions per ICU patient-day. (**C**) Distribution of ICU-days spent in each clinical state per patient. Y-axis is discontinuous to accommodate all data points.

**Figure 5.**
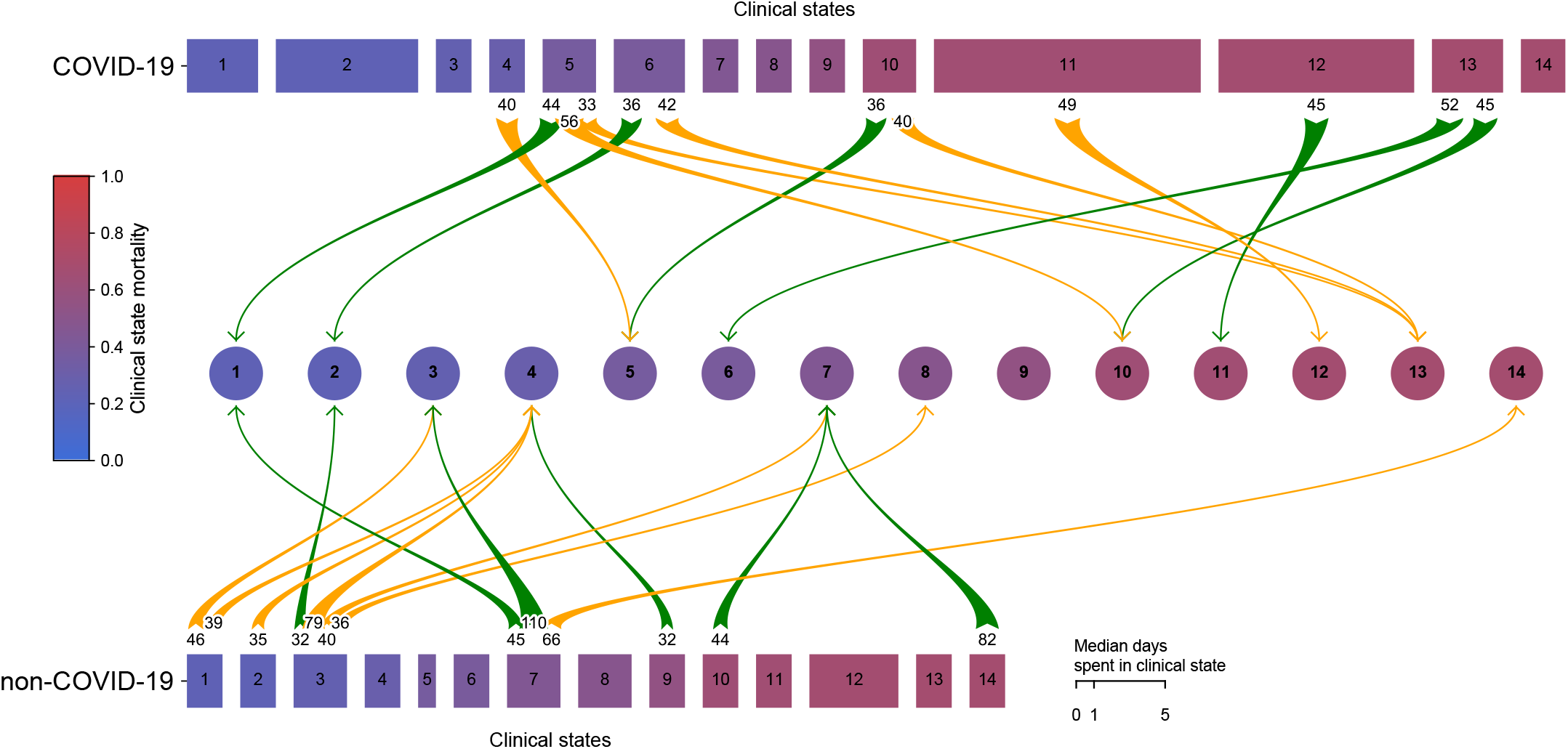
Patients with SARS-CoV-2 pneumonia have a longer length of stay and fewer transitions between clinical states per day compared to patients with non-COVID-19-related respiratory failure. Clinical states are ordered and numbered (1-14) by their associated mortality rate (blue to red). Rectangle width reflects median days per clinical state. Transitions marked by green arrows are to a more favorable (lower mortality) clinical state; yellow arrows mark transitions to a less favorable (higher mortality) clinical state. Numbers at the arrow bases represent the number of transitions between the two clinical states connected by the arrow. Only transitions that occurred more than 30 times are shown.

CarpeDiem *reveals unresolving VAP as a driver of poor outcomes in patients with severe pneumonia*. Overall, 35.4% of patients in the cohort developed at least one episode of VAP during their ICU stay (24.8% among patients without COVID-19 compared to 57.4% among patients with COVID-19, p<0.001) (Figure 6A). 8.5% of patients in the cohort experienced more than one episode of VAP (3.3% among patients without COVID-19 compared to 19.5% among patients with COVID-19, p<0.001) (Figure 6B). Mortality in patients with VAP has been reported to increase substantially with each ensuing episode, approaching 100% in patients with three or more episodes.^31^ In contrast, we found that the mortality associated with a single VAP episode did not differ from the mortality associated with multiple VAP episodes (48.6% with a single episode, 52.5% with two episodes, 50.0% with three episodes; p=not significant) (Figure 6C), suggesting that cure can be achieved even in patients with multiple VAP episodes. Nevertheless, the relatively small number of patients with multiple VAP episodes limits power to detect small differences (Figure 6D).

**Figure 6.**
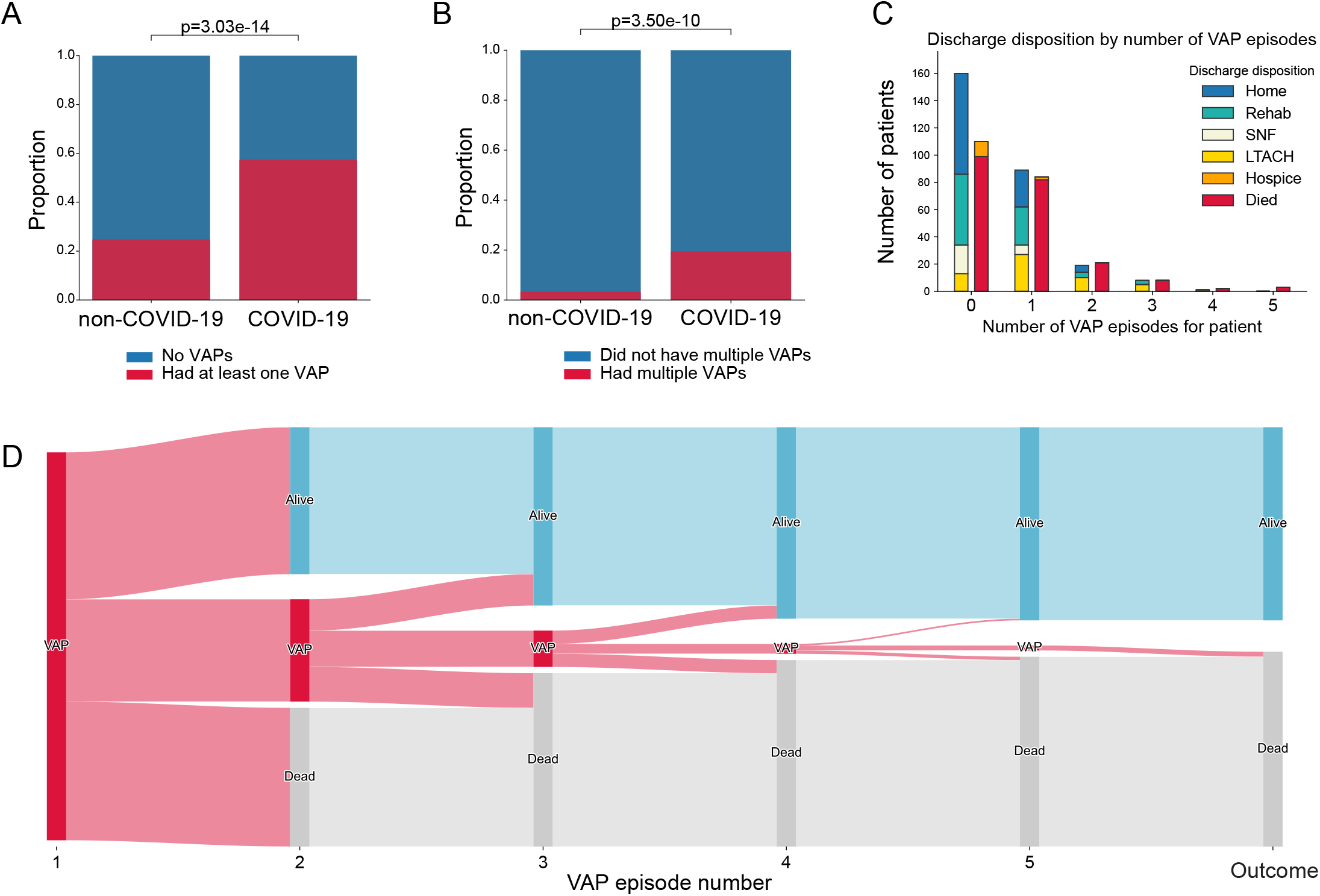
Patients with COVID-19 experience more VAPs compared to patients without COVID-19. **(A)** Proportion of patients with or without COVID-19 having at least one VAP. (**B**) Proportion of patients with or without COVID-19 having more than one VAP. (**C**) Outcomes for patients experiencing different numbers of VAP episodes. Outcomes are displayed in two columns: the first column aggregates favorable discharge dispositions (Home, Rehab, SNF, LTACH), the second column aggregates unfavorable discharge dispositions (Hospice, Died). (**D**) Sankey diagram of VAP episodes and outcomes for each VAP episode. SNF = skilled nursing facility, LTACH = long-term acute care hospital.

Mortality was not statistically different in patients who developed VAP compared to those who did not (Figure 7A). To further explore the association between VAP and ICU outcomes, we used the clinical adjudication results from the SCRIPT study to compare patients with successful treatment of VAP (cured) with those who experienced an indeterminate outcome or unsuccessful treatment (not cured). Using this analysis applied to patients who had only a single VAP episode, we found that mortality was lowest among patients with successful treatment (cured), intermediate among those with an indeterminate outcome, and highest among those with unsuccessful treatment (not cured) (Figure 7B). We also observed a similar pattern among the subset of patients with COVID-19 (Supplemental Figure 8A). Patients with COVID-19 experienced longer durations of VAP episodes (Figure 7C). Unresolving VAP episodes (those with an indeterminate outcome or that were not cured) were of longer duration than cured episodes (Figure 7D). Since survival is included in our definition of successful VAP treatment, we performed a sensitivity analysis on VAP episodes experienced by patients who survived for at least 14 days following VAP diagnosis. Even among this group biased toward better outcomes, we found that unresolving VAP was associated with increased mortality (Supplemental Figure 8B).

**Figure 7.**
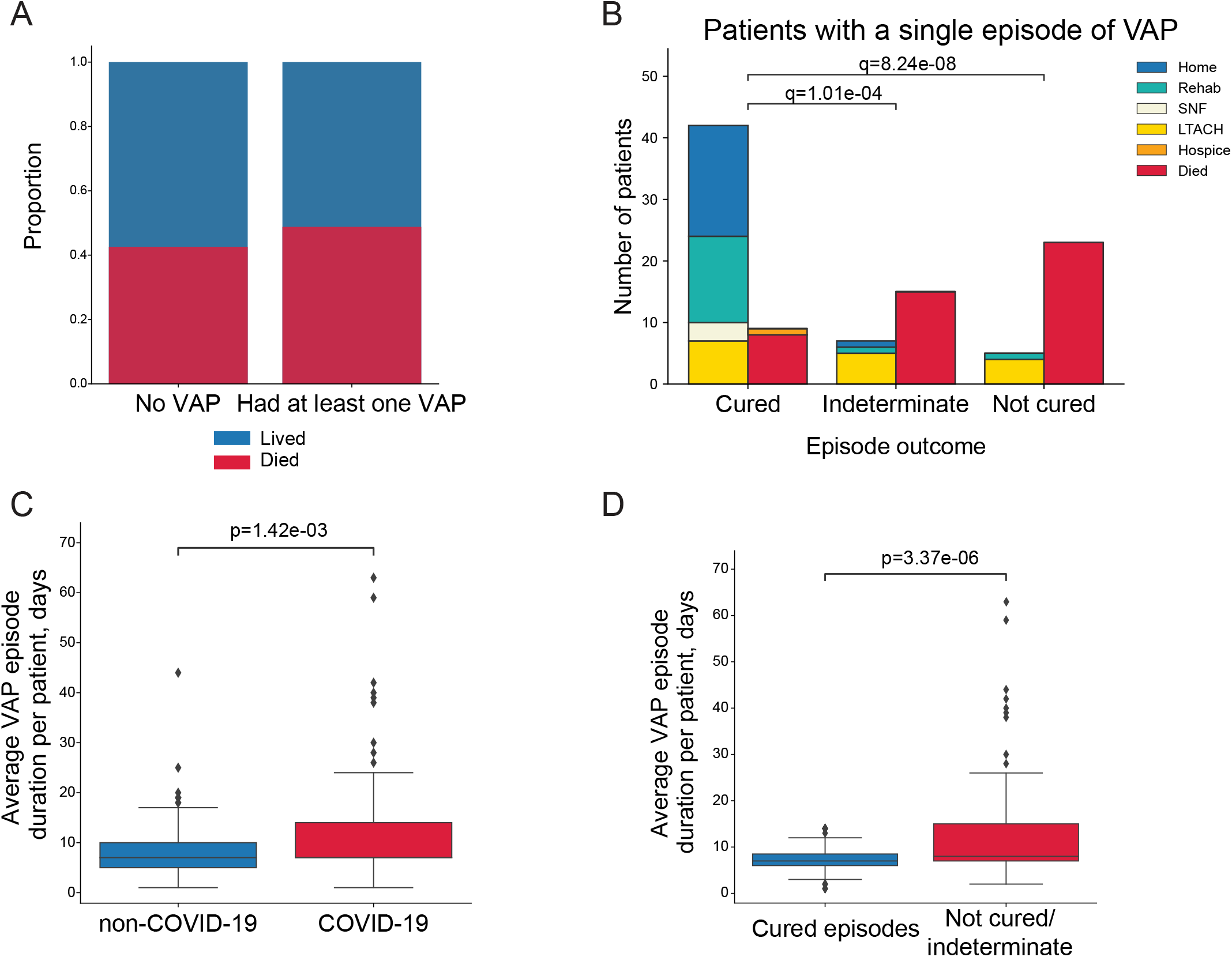
Unresolving VAP is associated with worse outcomes. (**A**) Mortality associated with having at least one episode of VAP. (**B**) Outcomes for patients who experienced one episode of VAP that was cured, of indeterminate cure status, or not cured by 14 days following diagnosis. Outcomes are displayed in two columns: the first column aggregates favorable discharge dispositions (Home, Rehab, SNF, LTACH), the second column aggregates unfavorable discharge dispositions (Hospice, Died). (**C**) VAP episode duration in patients with COVID-19 compared to patients without COVID-19. (**D**) VAP episode duration in patients who were cured or not cured/indeterminate cure status. SNF = skilled nursing facility, LTACH = long-term acute care hospital.

We then used the transition analyses provided by *CarpeDiem* to test whether unresolving VAP was associated with a subsequent trajectory toward progressively unfavorable clinical states. Successful treatment of VAP was associated with an increased likelihood of favorable subsequent transitions (Figure 8A). In contrast, indeterminate episodes demonstrated a flat trajectory (Figure 8B). Not cured episodes were associated with an increased risk of unfavorable subsequent transitions (Figure 8C, Supplemental Figure 9A-B).

**Figure 8.**
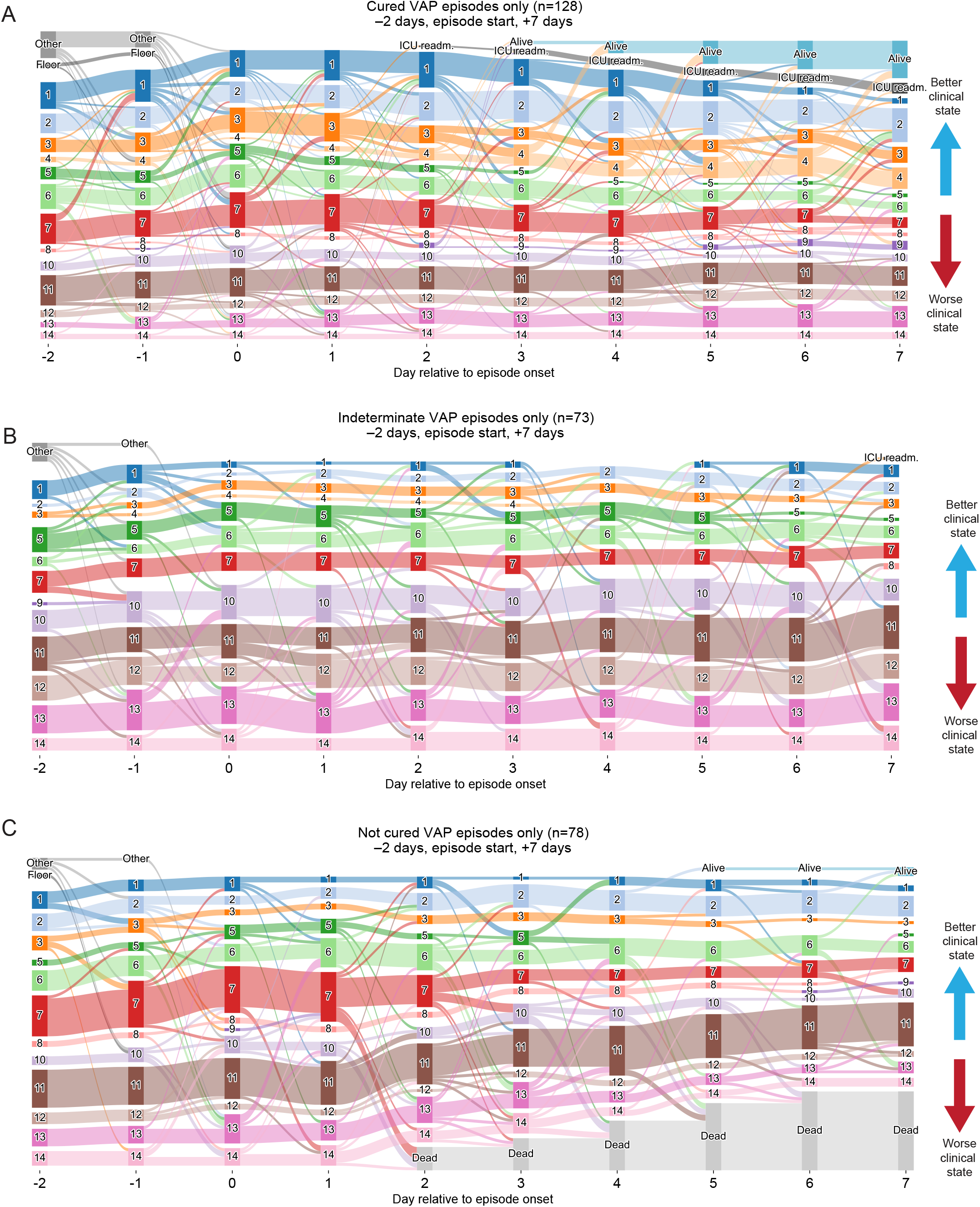
Trajectory analysis reveals that unresolving VAP is associated with transitions to progressively unfavorable clinical states. On these Sankey diagrams, day 0 represents the day that a BAL procedure was performed to evaluate suspected VAP adjudicated as (**A**) cured, (**B**) indeterminate, or (**C**) not cured. More favorable (lower mortality) clinical states are at the top of the graphs, with leaving the ICU alive being the highest, and less favorable (higher mortality) clinical states are at the bottom, with death being the lowest. Graphs start at two days prior to episode onset; patients who were not in our ICU are labeled as ‘Other’ (external transfers, home chronically ventilated patients) or ‘Floor’ (within 48 hours of extubation, or chronically ventilated patients).

To assess whether the same associations could be revealed independent of the *CarpeDiem* approach, we added flags denoting the development of VAP and its outcome to a standard model of ICU mortality prediction based on clinical parameters measured early (in the first two days) of ICU admission. Using gradient boosting, we found only a nominal increase in the predictive ability of early clinical parameters with addition of the VAP flags (Supplemental Figure 10A-C). These dissonant findings are possibly explained by the disconnect between clinical parameters measured early in a clinical course and the fact that VAP, by definition, occurs later in an ICU stay.

## Discussion

The ICU course of patients with severe SARS-CoV-2 pneumonia is more than twice as long as the duration in patients with pneumonia caused by other pathogens.^3–8^ Despite significantly longer durations of critical illness, mortality in patients with COVID-19 is similar to patients with other etiologies of pneumonia and respiratory failure.^4–6,8^ This finding is surprising, as ICU LOS and duration of mechanical ventilation have historically been associated with poor outcomes and have been used as intermediate endpoints in clinical trials.^32^ We and others have reported unexpectedly high rates of VAP complicating the ICU course of patients with SARS-CoV-2 pneumonia.^2,9^ Accordingly, we conceptualized a model in which the discordance between ICU LOS and mortality in patients with severe SARS-CoV-2 pneumonia results from a low mortality attributable to the primary viral pneumonia that is offset by an increased risk of mortality from unresolving VAP or other ICU complications.

To test the hypothesis that unresolving VAP explains the apparent disconnect between ICU LOS and mortality, we developed a machine learning approach, *CarpeDiem*, that uses clinical data extracted from the EHR to discretize days in the ICU into clinical states. We combined *CarpeDiem* with expert adjudication of pneumonia episodes and outcomes based on prospectively defined criteria. Our analysis revealed that the long ICU LOS in patients with SARS-CoV-2 pneumonia relative to those with pneumonia secondary to other pathogens is attributable to relatively long stays in clinical states primarily characterized by severe respiratory failure. When normalized for ICU LOS, patients with SARS-CoV-2 pneumonia experience fewer transitions between clinical states relative to other patients over the course of their ICU stay. In patients with SARS-CoV-2 pneumonia and others in the cohort, unresolving VAP was associated with transitions toward progressively unfavorable clinical states and increased hospital mortality compared with successfully treated VAP.

Guidelines adopted by professional societies recommend a host of interventions to prevent and treat known or suspected VAP in patients requiring mechanical ventilation, implicitly acknowledging the importance of VAP in determining outcomes.^16,33^ Proving the association between VAP and outcomes, including the mortality directly attributable to VAP, has been challenging because the risk of VAP is directly related to the duration of mechanical ventilation. Furthermore, the same patient can develop more than one episode of VAP, and non-specific and poorly sensitive techniques are often used to diagnose VAP episodes and adjudicate VAP outcomes.^16,34,35^ To address this challenge, we based *CarpeDiem* on the common practice of daily ICU rounds during which ICU teams use clinical data to assess each patient’s trajectory and develop or adjust a plan of care.^36^ *CarpeDiem* clustered ICU patient-days into common clinical states associated with differential mortality, driven by differences in the number and severity of organ failures.^26,28,30^ Almost all of the patients in our cohort underwent one or more transitions between these clinical states. We defined these transitions as favorable or unfavorable based on the mortality associated with the clinical states, providing an intermediate outcome to assess the impact of intercurrent ICU interventions or complications on outcome. We combined *CarpeDiem* with a rigorous, BAL-based approach to adjudicate episodes of VAP and their outcomes based on state-of-the art analysis of BAL fluid with multiplex PCR and quantitative culture.^2,13,37^ This approach revealed a strong association between unresolving episodes of VAP and mortality. In contrast, successfully treated VAP was associated with improved outcomes in all patients with severe respiratory failure, including due to SARS-CoV-2.

The importance of VAP as a driver of mortality in patients with COVID-19 has been underestimated, likely because bronchoscopic sampling has been uncommon during the pandemic, use of antibiotics is ubiquitous, and clinical criteria and biomarkers do not accurately distinguish between primary SARS-CoV-2 pneumonia and secondary bacterial pneumonia.^38^ For example, only one episode of secondary pneumonia was reported in the 403 patients included in the REMAP-CAP trial of hydrocortisone for COVID-19, and no episodes were reported in the 6,425 patients included in the RECOVERY trial of dexamethasone therapy for COVID-19.^3,39^ If unresolving episodes of VAP rather than the primary viral pneumonia contribute to mortality in a substantial fraction of patients with severe SARS-CoV-2 pneumonia, it might explain why therapies that attenuate the host response (e.g., corticosteroids, IL-6 receptor antagonists, JAK2 inhibitors, and CRAC channel inhibitors) are more effective when administered early in the clinical course, before patients are intubated and at risk for VAP.^3,40–43^

The *CarpeDiem* approach also provided insight into the long length of stay in patients with SARS-CoV-2 pneumonia. We found this long length of stay was explained by prolonged stays in clinical states characterized by severe respiratory failure. Transitions between clinical states per ICU-day were lower in patients with SARS-CoV-2 pneumonia compared to the rest of the cohort. These findings are consistent with models of SARS-CoV-2 pneumonia pathobiology in which the sparse expression of ACE2, the receptor for SARS-CoV-2, in the lung epithelium results in a slowly progressive but spatially localized infection that unfolds over days to weeks.^8,44,45^ It is also consistent with autopsy studies that have used ultra-sensitive techniques, including single-cell RNA-sequencing and sm-FISH, to demonstrate that SARS-CoV-2 infection is largely limited to the airway and alveolar epithelium and alveolar macrophages.^46,47^

Our study has important limitations. First, as ours is an observational study, we cannot exclude unmeasured confounders that link unresolving VAP to poor outcomes. Second, we used state-of-the art microbiological analysis of distal lung samples to diagnose VAP, and we have shown that clinicians in our center use this information to optimize, narrow, or discontinue antibiotic therapy,^2^ minimizing the harmful effects of inappropriate or excessive antibiotic therapy.^48^ Hence, the reasons underlying the failure of appropriate antimicrobial therapy in some patients cannot be determined. Further studies of the host response, microbe, and microbiome may provide insights into these mechanisms. Third, *CarpeDiem* uses a limited number of parameters to define clinical states, potentially neglecting important determinants of outcome and information that might be found in missing data (e.g., reduced monitoring and ordering of laboratory tests as patients improve or move toward comfort-focused care). Similarly, intermittently measured biomarkers associated with outcomes, for example those used by Calfee et al. to define hyper- and hypo-inflammatory states in the ICU, are incompletely represented in *CarpeDiem*.^49^ Future iterations of the tool can incorporate these data with a goal of improving the association between clinical states and mortality. To this end, we have made de-identified data from the SCRIPT dataset, as well as detailed code, freely available to the research community.

### Data availability

The dataset will be available on PhysioNet.

The demonstration browser is available at: https://nupulmonary.org/carpediem Code is available at: https://github.com/NUSCRIPT/carpediem

## Supporting information

Supplemental File 2: NU SCRIPT Study Investigators

Supplemental File 1: Pneumonia Adjudication

## Data Availability

Data will be available online on PhysioNet. Code is available at https://github.com/NUSCRIPT/carpediem. Data browser is available at https://nupulmonary.org/carpediem/.

## Author contributions

CAG, NSM, TS, GRSB, RGW, AVM, BDS conceived, designed, interpreted results, and wrote/edited the manuscript.

CAG, NSM, TS performed programming and analysis.

CAG, CP, JMW, JMK, RGW, BDS performed clinical pneumonia episode adjudication. HKD and AD enrolled patients and compiled clinical adjudication data.

AP, PN, RAG, CAG, MK, LR, DS, JS compiled the dataset.

CAG, NSM, TS, MK, LR, DS, JS have directly accessed and verified the underlying data reported in the manuscript.

All authors confirm that they had full access to all the data in the study and accept responsibility to submit for publication. All authors read and approved the final draft of the manuscript.

## Acknowledgements

The authors would like to thank Malte Luecken and Neal Ravindra for valuable discussion. SCRIPT is supported by NIH/NIAID U19AI135964.

This study was also supported by the Canning Thoracic Institute of Northwestern Medicine. CAG is supported by NIH/NHLBI Training Grant T32HL076139 and F32HL162377.

TS is supported by K99AG068544.

GRSB was supported by NIH grants U19AI135964, P01AG049665, R01HL147575, P01HL071643,

R01HL154686, Veterans Affairs grant I01CX001777, a grant from the Chicago Biomedical Consortium and a Northwestern University Dixon Translational Science Award.

RGW is supported by NIH grants U19AI135964, U01TR003528, P01HL154998, R01HL14988, R01LM013337 AVM is supported by NIH awards U19AI135964, P01AG049665, R21AG075423, R01HL158139, R01HL153312, P01HL154998.

BDS is supported by NIH awards R01HL149883, R01HL153122, P01HL154998, P01AG049665, and U19AI135964.

## Works Cited

1 Morens DM, Taubenberger JK, Fauci AS. Predominant role of bacterial pneumonia as a cause of death in pandemic influenza: implications for pandemic influenza preparedness. J Infect Dis 2008; 198: 962–70.

2 Pickens CO, Gao CA, Cuttica MJ, et al. Bacterial Superinfection Pneumonia in Patients Mechanically Ventilated for COVID-19 Pneumonia. Am J Respir Crit Care Med 2021; 204: 921–32.

3 RECOVERY Collaborative Group, Horby P, Lim WS, et al. Dexamethasone in Hospitalized Patients with Covid-19 - Preliminary Report. N Engl J Med 2020; published online July 17. DOI:10.1056/NEJMoa2021436.

4 Ferrando C, Suarez-Sipmann F, Mellado-Artigas R, et al. Clinical features, ventilatory management, and outcome of ARDS caused by COVID-19 are similar to other causes of ARDS. Intensive Care Med 2020; 46: 2200–11.

5 Gamberini L, Tonetti T, Spadaro S, et al. Factors influencing liberation from mechanical ventilation in coronavirus disease 2019: multicenter observational study in fifteen Italian ICUs. J Intensive Care Med 2020; 8: 80.

6 Roedl K, Jarczak D, Thasler L, et al. Mechanical ventilation and mortality among 223 critically ill patients with coronavirus disease 2019: A multicentric study in Germany. Aust Crit Care 2020; published online Oct 27. DOI:10.1016/j.aucc.2020.10.009.

7 Guan W-J, Ni Z-Y, Hu Y, et al. Clinical Characteristics of Coronavirus Disease 2019 in China. N Engl J Med 2020; 382: 1708–20.

8 Grant RA, Morales-Nebreda L, Markov NS, et al. Circuits between infected macrophages and T cells in SARS-CoV-2 pneumonia. Nature 2021; published online Jan 11. DOI:10.1038/s41586-020-03148-w.

9 Vacheron C-H, Lepape A, Savey A, et al. Attributable Mortality of Ventilator-associated Pneumonia Among COVID-19 Patients. Am J Respir Crit Care Med 2022; published online May 10. DOI:10.1164/rccm.202202-0357OC.

10 Viglianti EM, Bagshaw SM, Bellomo R, et al. Late vasopressor administration in patients in the ICU: A retrospective cohort study. Chest 2020; 158: 571–8.

11 Viglianti EM, Kramer R, Admon AJ, et al. Late organ failures in patients with prolonged intensive care unit stays. J Crit Care 2018; 46: 55–7.

12 Iwashyna TJ, Hodgson CL, Pilcher D, et al. Timing of onset and burden of persistent critical illness in Australia and New Zealand: a retrospective, population-based, observational study. Lancet Respir Med 2016; 4: 566–73.

13 Pickens CI, Wunderink RG. Principles and Practice of Antibiotic Stewardship in the ICU. Chest 2019; 156: 163–71.

14 Walter JM, Helmin KA, Abdala-Valencia H, Wunderink RG, Singer BD. Multidimensional assessment of alveolar T cells in critically ill patients. JCI Insight 2018; 3. DOI:10.1172/jci.insight.123287.

15 Walter JM, Ren Z, Yacoub T, et al. Multidimensional Assessment of the Host Response in Mechanically Ventilated Patients with Suspected Pneumonia. Am J Respir Crit Care Med 2019; 199: 1225–37.

16 Torres A, Niederman MS, Chastre J, et al. International ERS/ESICM/ESCMID/ALAT guidelines for the management of hospital-acquired pneumonia and ventilator-associated pneumonia: Guidelines for the management of hospital-acquired pneumonia (HAP)/ventilator-associated pneumonia (VAP) of the European Respiratory Society (ERS), European Society of Intensive Care Medicine (ESICM), European Society of Clinical Microbiology and Infectious Diseases (ESCMID) and Asociación Latinoamericana del Tórax (ALAT). Eur Respir J 2017; 50. DOI:10.1183/13993003.00582-2017.

17 Weiss CH, Moazed F, DiBardino D, Swaroop M, Wunderink RG. Bronchoalveolar lavage amylase is associated with risk factors for aspiration and predicts bacterial pneumonia. Crit Care Med 2013; 41: 765–73.

18 Starren JB, Winter AQ, Lloyd-Jones DM. Enabling a Learning Health System through a Unified Enterprise Data Warehouse: The Experience of the Northwestern University Clinical and Translational Sciences (NUCATS) Institute. Clin Transl Sci 2015; 8: 269–71.

19 Ward JH. Hierarchical Grouping to Optimize an Objective Function. Paperpile. https://paperpile.com/app/p/9d74dee9-431f-03b6-a0f1-5be2b24b7094 (accessed Sept 19, 2022).

20 Dixon JK. Pattern Recognition with Partly Missing Data. IEEE Trans Syst Man Cybern 1979; 9: 617–21.

21 Pedregosa F, Varoquaux G, Gramfort A, et al. Scikit-learn: Machine Learning in Python. J Mach Learn Res 2011; 12: 2825–30.

22 Johnson A, Bulgarelli L, Pollard T, Horng S, Celi LA, Mark R. MIMIC-IV. 2022. DOI:10.13026/7VCR-E114.

23 Johnson AE, Stone DJ, Celi LA, Pollard TJ. The MIMIC Code Repository: enabling reproducibility in critical care research. J Am Med Inform Assoc 2018; 25: 32–9.

24 Chen T, Guestrin C. XGBoost: A Scalable Tree Boosting System. In: Proceedings of the 22nd ACM SIGKDD International Conference on Knowledge Discovery and Data Mining. New York, NY, USA: Association for Computing Machinery, 2016: 785–94.

25 Goldberger AL, Amaral LA, Glass L, et al. PhysioBank, PhysioToolkit, and PhysioNet: components of a new research resource for complex physiologic signals. Circulation 2000; 101: E215–20.

26 Zimmerman JE, Kramer AA, McNair DS, Malila FM. Acute Physiology and Chronic Health Evaluation (APACHE) IV: hospital mortality assessment for today’s critically ill patients. Crit Care Med 2006; 34: 1297–310.

27 Vincent JL, Moreno R, Takala J, et al. The SOFA (Sepsis-related Organ Failure Assessment) score to describe organ dysfunction/failure. On behalf of the Working Group on Sepsis-Related Problems of the European Society of Intensive Care Medicine. Intensive Care Med 1996; 22: 707–10.

28 Vincent JL, de Mendonça A, Cantraine F, et al. Use of the SOFA score to assess the incidence of organ dysfunction/failure in intensive care units: results of a multicenter, prospective study. Working group on “sepsis-related problems” of the European Society of Intensive Care Medicine. Crit Care Med 1998; 26: 1793–800.

29 Zimmerman JE, Kramer AA, McNair DS, Malila FM, Shaffer VL. Intensive care unit length of stay: Benchmarking based on Acute Physiology and Chronic Health Evaluation (APACHE) IV. Crit Care Med 2006; 34: 2517–29.

30 Ferreira FL, Bota DP, Bross A, Mélot C, Vincent JL. Serial evaluation of the SOFA score to predict outcome in critically ill patients. JAMA 2001; 286: 1754–8.

31 Silver DR, Cohen IL, Weinberg PF. Recurrent Pseudomonas aeruginosa pneumonia in an intensive care unit. Chest 1992; 101. DOI:10.1378/chest.101.1.194.

32 Nelson JE, Cox CE, Hope AA, Carson SS. Chronic critical illness. Am J Respir Crit Care Med 2010; 182: 446–54.

33 Kalil AC, Metersky ML, Klompas M, et al. Management of Adults With Hospital-acquired and Ventilator-associated Pneumonia: 2016 Clinical Practice Guidelines by the Infectious Diseases Society of America and the American Thoracic Society. Clin Infect Dis 2016; 63: e61–111.

34 Weiss E, Zahar JR, Alder J, et al. Elaboration of consensus clinical endpoints to evaluate antimicrobial treatment efficacy in future HABP/VABP clinical trials. Clin Infect Dis 2019. DOI:10.1093/cid/ciz093.

35 Talbot GH, Powers JH, Hoffmann SC, et al. Developing outcomes assessments as endpoints for registrational clinical trials of antibacterial drugs: 2015 update from the Biomarkers Consortium of the Foundation for the National Institutes of Health. Clin Infect Dis 2016; 62: 603–7.

36 Lighthall GK, Vazquez-Guillamet C. Understanding Decision Making in Critical Care. Clin Med Res 2015; 13: 156–68.

37 Pickens C, Wunderink RG, Qi C, et al. A multiplex polymerase chain reaction assay for antibiotic stewardship in suspected pneumonia. Diagn Microbiol Infect Dis 2020; 98: 115179.

38 Gao CA, Bailey JI, Walter JM, et al. Bronchoscopy on Intubated COVID-19 Patients is Associated with Low Infectious Risk to Operators. Ann Am Thorac Soc 2021; published online Jan 15. DOI:10.1513/AnnalsATS.202009-1225RL.

39 Tomazini BM, Maia IS, Cavalcanti AB, et al. Effect of Dexamethasone on Days Alive and Ventilator-Free in Patients With Moderate or Severe Acute Respiratory Distress Syndrome and COVID-19: The CoDEX Randomized Clinical Trial. JAMA 2020; 324: 1307–16.

40 Angus DC, Derde L, Al-Beidh F, et al. Effect of Hydrocortisone on Mortality and Organ Support in Patients With Severe COVID-19: The REMAP-CAP COVID-19 Corticosteroid Domain Randomized Clinical Trial. JAMA 2020; 324: 1317–29.

41 Angriman F, Ferreyro BL, Burry L, et al. Interleukin-6 receptor blockade in patients with COVID-19: placing clinical trials into context. Lancet Respir Med 2021; published online April 27. DOI:10.1016/S2213-2600(21)00139-9.

42 Kalil AC, Patterson TF, Mehta AK, et al. Baricitinib plus Remdesivir for Hospitalized Adults with Covid-19. N Engl J Med 2020; published online Dec 11. DOI:10.1056/NEJMoa2031994.

43 Bruen C, Al-Saadi M, Michelson EA, et al. Auxora vs. placebo for the treatment of patients with severe COVID-19 pneumonia: a randomized-controlled clinical trial. Crit Care 2022; 26: 101.

44 Muus C, Luecken MD, Eraslan G, et al. Single-cell meta-analysis of SARS-CoV-2 entry genes across tissues and demographics. Nat Med 2021; 27: 546–59.

45 Budinger GRS, Misharin AV, Ridge KM, Singer BD, Wunderink RG. Distinctive features of severe SARS-CoV-2 pneumonia. J Clin Invest 2021; 131. DOI:10.1172/JCI149412.

46 Delorey TM, Ziegler CGK, Heimberg G, et al. COVID-19 tissue atlases reveal SARS-CoV-2 pathology and cellular targets. Nature 2021; published online April 29. DOI:10.1038/s41586-021-03570-8.

47 Bharat A, Querrey M, Markov NS, et al. Lung transplantation for patients with severe COVID-19. Sci Transl Med 2020; 12. DOI:10.1126/scitranslmed.abe4282.

48 Kollef MH, Chastre J, Clavel M, et al. A randomized trial of 7-day doripenem versus 10-day imipenem-cilastatin for ventilator-associated pneumonia. Crit Care 2012; 16: R218.

49 Calfee CS, Delucchi K, Parsons PE, et al. Subphenotypes in acute respiratory distress syndrome: latent class analysis of data from two randomised controlled trials. Lancet Respir Med 2014; 2: 611–20.

