## Supplemental File 1: Pneumonia Adjudication for "A machine learning approach identifies unresolving secondary pneumonia as a contributor to mortality in patients with severe pneumonia, including COVID-19"

### Investigator SCRIPT Pneumonia and Viral Episode Assessment and Outcome Evaluation

Study ID: ______________ Reviewer ○ 1 ○ 2 ○ 3 ○ CR: ________ (Date)

| ○ N/A – go to page 2 | **Episode Category Assessment** |
|---|---|
| Is this for the initial pneumonia episode?<br><br>**Date of BAL in REDCap**: _______________ | ○ Yes, this is the initial pneumonia episode (implies > 1 episodes)<br>○ Yes, this is the initial and also final pneumonia episode<br>○ No, multiple episodes |
| If not initial, is this the final pneumonia episode? | ○ Yes ○ No ○ N/A |
| Patient Category:<br>○ Clinical CAP (not hospitalized within the last 7 days)<br>○ Clinical HAP (current admission >48 hours or discharged from a healthcare facility within the last 7 days where admission >24 hours)<br>○ Clinical VAP (on ventilator >48 hours or reintubated < 24 hours from extubation)<br>○ Non-pneumonia control*<br>○ Infection [a]<br>○ Known Condition [b]<br>○ Unknown | *If non-pneumonia control, cause of infiltrate:<br>○ ARDS<br>○ Aspiration<br>○ Atelectasis<br>○ Fibrosis<br>○ Fluid overload<br>○ Heart failure/pulmonary edema<br>○ Pleural effusion<br>○ Pulmonary hemorrhage<br>○ Other: __________________<br>○ Unknown |
| [a] *If infection:*<br>○ Cholangitis/cholecystitis<br>○ Colitis<br>○ Other intra-abdominal<br>○ Line infection<br>○ Tracheobronchitis<br>○ Urinary tract<br>○ Wound/skin<br>○ Other: _____________________ | [b] *If known condition, cause of fever/leukocytosis:*<br>○ Aspiration<br>○ Atelectasis<br>○ Drug fever<br>○ Pancreatitis<br>○ Other: _____________________<br>○ None<br>○ Unknown |
| **STOP here for non-pneumonia control** | |
| Choose one: (from Episode Category Assessment)<br>○ Viral* only<br>○ Bacterial/Viral* Co-Infection<br>○ Bacterial/Etiology defined<br>○ Culture-negative** (%PMNs ≥ 50%)<br>○ Culture-negative*** (%PMNs < 50%)<br>○ Indeterminate | ******If viral, select virus type: (select all that apply)*<br>☐ Influenza<br>☐ SARS-CoV-2<br>☐ Other:____________ |
| **Demographics page immune status is** ○ Immunocompromised ○ NON-Immunocompromised | |
| ***If culture-negative (%PMNs ≥ 50%):*<br>○ Immunocompromised<br>○ Nonimmunocompromised | ****If culture-negative (%PMNs < 50%):*<br>○ Neutropenic (ANC < 500/uL)<br>○ Immunocompromised<br>○ Nonimmunocompromised |

| **If Viral Only** ○ N/A | |
|---|---|
| Were appropriate antivirals administered? | ○ Yes* ○ No ○ N/A – no appropriate antivirals |
| **If yes**, select antiviral: (select all that apply)* | ☐ Influenza – oseltamivir<br>☐ Influenza-baloxavir<br>☐ Influenza-peramivir<br>☐ Influenza-other: ________________________<br>☐ SARS-CoV-2-remdesivir<br>☐ SARS-Cov-2-other: ________________________<br>☐ Adenovirus-cidofovir<br>☐ Herpes or Varicella-acyclovir<br>☐ RSV-ribavirin only if immunocompromised<br>☐ Other (please specify): ________________________ |
| Were empirical antibiotics for this episode started prior to BAL? | ○ Yes* ○ No |
| *If **yes**, number of days **prior** to BAL collection?* | ○ ≤ 48 hours before BAL<br>○ > 48 hours before BAL |
| Were empirical antibiotics **started/continued** after BAL collection? | ○ Yes* ○ No |
| *If **yes**, select number of days antibiotics were given:* | ○ < 72 hours after BAL<br>○ 3-7 days after BAL<br>○ 8-10 days after BAL<br>○ 11-14 days after BAL<br>○ ≥ 15 days after BAL |
| **Viral Pneumonia Clinical Impression** | |
| Was viral retesting performed? | ○ Yes*<br>○ No (go to Global Viral Clinical Impression) |
| *If **yes**, PCR cleared?* | ○ Yes by BAL<br>Date of 1st negative test with no subsequent positive: ________________<br>○ Yes by NP/OP (no BAL available or subsequently completed)<br>Date of 1st persistent negative test: ____________<br>○ Other viral procedure: __________________<br>Date of repeat test: ____________________<br>○ No<br>Date of last positive test: ________________ |
| Global Viral Clinical Impression:<br>(*Go to PCT after*) | **Pneumonia (Check best answer)**<br>○ Cure<br>○ Persistence (repeat positive PCR until death, transfer to LTAC, needs time interval from last testing [i.e. positive within 7 days])<br>○ *Superinfection pneumonia (N/A if single episode only)*<br>○ Indeterminate (NP positive but BAL clear, no recent PCR prior to death or transfer to other facility)<br><br>**Other: (if applicable, all that apply)**<br>☐ Extrapulmonary infection (Single Viral) ***(see extra question)**** |

If ***extrapulmonary infection,** choose site (**check all that apply**) and corresponding documentation**:**

| **Site *(check all that apply)*** | **Documentation *(check most appropriate – only one)*** Definitive with positive culture | Definitive without culture proof | Radiologic/imaging only | Clinical suspicion only |
|---|---|---|---|---|
| ☐ Cholangitis/cholecystitis | ○ | ○ | ○ | ○ |
| ☐ Colitis /C. diff | ○ | ○ | ○ | ○ |
| ☐ Other intra-abdominal (SBP, abcess, etc.) | ○ | ○ | ○ | ○ |
| ☐ Line infection | ○ | ○ | ○ | ○ |
| ☐ Tracheobronchitis | ○ | ○ | ○ | ○ |
| ☐ Urinary Tract | ○ | ○ | ○ | ○ |
| ☐ Wound/skin | ○ | ○ | ○ | ○ |
| ☐ Empyema (unrelated to the index pna) | ○ | ○ | ○ | ○ |
| ☐ Other (*please specify*): ____________________ | ○ | ○ | ○ | ○ |

| **If Bacterial/Viral Co-Infection** ○ N/A (Viral Only) | |
|---|---|
| Were appropriate antivirals administered? | ○ Yes* ○ No ○ N/A – no appropriate antivirals |
| **If yes*, select antiviral: (select all that apply)* | ☐ Influenza – oseltamivir<br>☐ Influenza-baloxavir<br>☐ Influenza-peramivir<br>☐ Influenza-other: _________________________<br>☐ SARS-CoV-2-remdesivir<br>☐ SARS-CoV-2-other: _________________________<br>☐ Adenovirus-cidofovir<br>☐ Herpes or Varicella-acyclovir<br>☐ RSV-ribavirin only if immunocompromised<br>☐ Other (please specify): _________________________ |
| Was viral retesting performed? | ○ Yes*<br>○ No (go to *If initial sample, has the patient…*) |
| **If yes*, PCR cleared?* | ○ Yes by BAL<br>Date of 1st negative test with no subsequent positive: _______________<br>○ Yes by NP/OP (no BAL available or subsequently completed)<br>Date of 1st persistent negative test: ___________<br>○ Other viral procedure: _________________<br>Date of repeat test: ____________________<br>○ No<br>Date of last positive test: _______________ |

| **Continue If Bacterial/Viral Co-Infection** or **Complete If Bacterial Only** ○ N/A (Viral only) | |
|---|---|
| If initial sample, has the patient been actively treated for pneumonia for **more** than 24 hours prior to sample collection? | ○ Yes* ○ No ○ N/A<br><br>**BAL Collection Date/Time:** ______________________<br>*(Obtain from EPIC - Microbiology)* |
| **Starting antibiotics for pneumonia:** _____________<br><br>**Start date for antibiotics for pneumonia:** _____________<br><br>**If* ***yes***, *how many calendar days has patient been treated? (Day BAL – Day abx started)* | __________________days ○ N/A |
| Appropriate initial antibiotics?<br>*(in the 72 hrs. preceding culture results)*<br><br>**Refer to biofire results if applicable* | ○ Yes ○ No ○ N/A – no bacterial pathogen |
| Were antibiotics for pneumonia discontinued on D 7-8? | ○ Yes*<br>○ No<br>○ N/A**(death/transfer to another institution)<br><br>**if* ***Yes****, finish following clinical impression and go to PCT*<br>***If* ***N/A****, skip "clinical impression" questions and go to PCT* |
| **Clinical Impression** | |
| Clinical Impression at **D7-8**:<br><br>**must be actively treated at this time point* | **Pneumonia (Check best answers)**<br>○ Cure<br>☐ Persistence *(check all that apply)*<br>☐ Positive Culture<br>☐ Positive PCR<br>☐ Abscess / Cavity<br>☐ Empyema<br>☐ Endocarditis<br>☐ Other: ________________________<br>☐ Superinfection pneumonia<br>☐ Indeterminate<br><br>**Other: (if applicable, all that apply)**<br>☐ Extrapulmonary infection ***(see extra question)****<br>☐ Prophylaxis only |

If ***extrapulmonary infection,** choose site (**check all that apply**) and corresponding documentation**:**

| | **Documentation** ***(check most appropriate – only one)*** | | | |
|---|---|---|---|---|
| **Site** ***(check all that apply)*** | Definitive with positive culture | Definitive without culture proof | Radiologic/imaging only | Clinical suspicion only |
| ☐ Cholangitis/cholecystitis | ○ | ○ | ○ | ○ |
| ☐ Colitis /C. diff | ○ | ○ | ○ | ○ |
| ☐ Other intra-abdominal (SBP, abcess, etc.) | ○ | ○ | ○ | ○ |
| ☐ Line infection | ○ | ○ | ○ | ○ |
| ☐ Tracheobronchitis | ○ | ○ | ○ | ○ |
| ☐ Urinary Tract | ○ | ○ | ○ | ○ |
| ☐ Wound/skin | ○ | ○ | ○ | ○ |
| ☐ Empyema (unrelated to the index pna) | ○ | ○ | ○ | ○ |
| ☐ Other (*please specify*): ____________________ | ○ | ○ | ○ | ○ |

| | |
|---|---|
| Were antibiotics for pneumonia discontinued on D10? | ○ Yes*<br>○ No<br>○ N/A**(death/transfer to another institution)<br><br>**if* ***Yes****, finish following clinical impression and go to PCT*<br>***If* ***N/A****, skip "clinical impression" questions and go to PCT* |
| Clinical Impression at **D10**:<br><br>**must be actively treated at this time point* | **Pneumonia (Check best answers)**<br>○ Cure<br>☐ Persistence *(check all that apply)*<br> ☐ Positive Culture<br> ☐ Positive PCR<br> ☐ Abscess / Cavity<br> ☐ Empyema<br> ☐ Endocarditis<br> ☐ Other: ______________________<br>☐ Superinfection pneumonia<br>☐ Indeterminate<br><br>**Other: (if applicable, all that apply)**<br>☐ Extrapulmonary infection ***(see extra question)****<br>☐ Prophylaxis only |

If ***extrapulmonary infection,** choose site (**check all that apply**) and corresponding documentation**:**

| **Site *(check all that apply)*** | **Documentation *(check most appropriate – only one)*** Definitive with positive culture | Definitive without culture proof | Radiologic/imaging only | Clinical suspicion only |
|---|---|---|---|---|
| ☐ Cholangitis/cholecystitis | ○ | ○ | ○ | ○ |
| ☐ Colitis /C. diff | ○ | ○ | ○ | ○ |
| ☐ Other intra-abdominal (SBP, abcess, etc.) | ○ | ○ | ○ | ○ |
| ☐ Line infection | ○ | ○ | ○ | ○ |
| ☐ Tracheobronchitis | ○ | ○ | ○ | ○ |
| ☐ Urinary Tract | ○ | ○ | ○ | ○ |
| ☐ Wound/skin | ○ | ○ | ○ | ○ |
| ☐ Empyema (unrelated to the index pna) | ○ | ○ | ○ | ○ |
| ☐ Other (*please specify*): ____________________ | ○ | ○ | ○ | ○ |

| | |
|---|---|
| Were antibiotics for pneumonia discontinued on D14? | ○ Yes*<br>○ No<br>○ N/A**(death/transfer to another institution)<br>**if **Yes**, finish following clinical impression and go to PCT*<br>***If **N/A**, skip "clinical impression" questions and go to PCT* |
| *If **no**, what date were antibiotics for pneumonia stopped?* | Stop date for antibiotics for PNA: ____________<br>○ Stop date unknown (patient was discharged on antibiotics for pneumonia) |
| Clinical Impression at **D14**:<br><br>**must be actively treated at this time point* | **Pneumonia (Check best answers)**<br>○ Cure<br>☐ Persistence *(check all that apply)*<br>☐ Positive Culture<br>☐ Positive PCR<br>☐ Abscess / Cavity<br>☐ Empyema<br>☐ Endocarditis<br>☐ Other: ______________________<br>☐ Superinfection pneumonia<br>☐ Indeterminate<br><br>**Other: (if applicable, all that apply)**<br>☐ Extrapulmonary infection ***(see extra question)****<br>☐ Prophylaxis only |

If ***extrapulmonary infection,** choose site (**check all that apply**) and corresponding documentation**:**

| | **Documentation** ***(check most appropriate – only one)*** | | | |
|---|---|---|---|---|
| **Site** ***(check all that apply)*** | Definitive with positive culture | Definitive without culture proof | Radiologic/imaging only | Clinical suspicion only |
| ☐ Cholangitis/cholecystitis | ○ | ○ | ○ | ○ |
| ☐ Colitis /C. diff | ○ | ○ | ○ | ○ |
| ☐ Other intra-abdominal (SBP, abcess, etc.) | ○ | ○ | ○ | ○ |
| ☐ Line infection | ○ | ○ | ○ | ○ |
| ☐ Tracheobronchitis | ○ | ○ | ○ | ○ |
| ☐ Urinary Tract | ○ | ○ | ○ | ○ |
| ☐ Wound/skin | ○ | ○ | ○ | ○ |
| ☐ Empyema (unrelated to the index pna) | ○ | ○ | ○ | ○ |
| ☐ Other (*please specify*): ____________________ | ○ | ○ | ○ | ○ |

**PROCALCITONIN: (within 72 hrs of BAL/NBBAL Collection) [All categories]**

| | |
|---|---|
| Are serial PCTs available?<br>*Note: minimum of 2 measurements within this treatment episode* | ○ Yes* ○ No |
| **If yes**, does the patient meet PCT criteria for cure?* | ○ Yes, (check criteria below) ○ No<br>☐ 80% change/decrease from baseline<br>☐ Absolute value of $\leq$ 0.5 ng/mL |

**IF FINAL EPISODE: [All categories]**

**Overall Global Clinical Cure** **please refer to guidelines*

Overall Success?

○ **Yes**

→ If Yes, all criteria? (see text box below)

- ○ Yes
- ○ No
  - → If No, was patient extubated **before** day 7?
    - ○ Yes
    - ○ No

1. survive duration of treatment
2. able to stop antibiotics for pneumonia
   a. Continuation of same antibiotics > 14 days but signs/symptoms of pneumonia stable (WBC, secretions, oxygenation, radiograph) or improving?
3. causative pathogen disappears from respiratory secretions or no further samples
   a. %PMNs in repeat BAL < 50% in non-neutropenic
4. clinical manifestations of pneumonia improve/resolve (e.g., fever, secretions, WBC, hypoxemia, septic shock)
5. ability to wean from vent or at least initiate SBTs
   a. Drop in minute ventilation and/or improvement in oxygenation
   b. Patient returned to pre-pneumonia ventilator/ECMO status or weaning from vent

○ **No**

→ Cause of failure:*

- ☐ Antibiotics for another indication in addition to suspected/proven PNA
- ☐ Persistence *(check all that apply)*
  - ☐ Positive Culture
  - ☐ Positive PCR
  - ☐ Abscess / Cavity
  - ☐ Empyema
  - ☐ Endocarditis
  - ☐ Other: ________________
- ☐ Persistent inflammation only
- ☐ Recurrence
- ☐ Superinfection pneumonia

* *Check best answers*
