## Supplemental Materials for "A machine learning approach identifies unresolving secondary pneumonia as a contributor to mortality in patients with severe pneumonia, including COVID-19"

**Supplemental Methods**

*Cohort and patient information*. Our cohort included mechanically-ventilated patients admitted to an intensive care unit (ICU) who were enrolled in the Successful Clinical Response in Pneumonia Therapy (SCRIPT) study from June 2018 to March 2022 at Northwestern Memorial Hospital (NMH). We analyzed only patient stays with a hospital discharge disposition and for which pneumonia episode cases had full clinical adjudication. We assigned patients to one of the four groups (non-pneumonia control, other pneumonia (bacterial), other viral pneumonia, or COVID-19) based on their initial SCRIPT enrollment bronchoalveolar lavage (BAL), as established by BAL pathogen culture tests and assessed by physician adjudicators (see below). Ages greater than 89 were grouped together per Safe Harbor guidelines[1] and reported as age 91, as has been done by other datasets.[2] Racial groups with fewer than five individuals were classified as 'Unknown or Not Reported' to protect patient anonymity. Patients who underwent lung transplantation for persistent respiratory failure were categorized as having died. In binarized outcomes, discharge to hospice or death were categorized as an outcome of 'Died,' whereas discharge to Home, Rehab, Skilled Nursing Facility, or Long-term Acute Care Hospital were categorized as 'Lived,' Supplemental Table 1 was generated using *tableone*.[3]

*ICU stay information.* Electronic Health Record (EHR) data are compiled by the Northwestern Medicine Enterprise Data Warehouse (EDW),[4] the primary data repository for clinical data at Northwestern. Approximately 150 data sources (including the main EHR system, Epic) are loaded into the EDW on a nightly basis. The data are primarily loaded using Microsoft technologies (Visual Studio, SSIS, etc.) and scheduled to load via the SQL Server Job Agent. Data engineers and architects on the EDW team then combine the data sources using custom SQL scripts, Visual Studio, SSIS, etc. to create datamarts. Analysts on the EDW team then work with the datamarts to create reports, dashboards, and extracts validated with clinician input.

Patients who had multiple ICU stays during the same hospitalization had their stays numbered consecutively but did not contribute data between their ICU stays. Multiple hospitalizations of the same patient are reported as separate patients, as SCRIPT enrollments are unique to each hospitalization. The dates of each per-day datapoint were removed and replaced with a day relative to the beginning of the ICU stay (initial day is day 1, with next day starting at 12:00 a.m.) for each ICU admission occurring during a hospitalization such that no dates are in the final dataset per Safe Harbor guidelines.[1]

*Clinical parameters.* We compiled the status of intubation, extracorporeal membrane oxygenation (ECMO), acute renal replacement therapy (hemodialysis [HD], and continuous renal replacement therapy [CRRT]), sedation parameters (Glasgow Coma Scale [GCS] subscores of eye opening, motor response, verbal response, and Richmond Agitation Sedation Scale [RASS]), lung injury ($PaO_2/FiO_2$ ratio, $PaO_2$, arterial pH, $PaCO_2$, PEEP, $FiO_2$, plateau pressure, lung compliance, and oxygen saturation), hemodynamics (norepinephrine rate in mcg/kg/min, a flag for norepinephrine, mean arterial pressure, systolic and diastolic blood pressure, lactic acid, hemoglobin, and bicarbonate), renal (creatinine, urine output, and aforementioned HD and CRRT flags), inflammatory markers (WBC count, neutrophil count, platelets, procalcitonin, C-reactive protein, D-dimer, lactate dehydrogenase, ferritin, bilirubin, albumin, and lymphocytes), vital signs (temperature, heart rate, respiratory rate), and ventilator instability (number of ventilator respiratory rate changes, positive end-expiratory pressure (PEEP) changes, and $FiO_2$ changes per day). When multiple measurements were available for the same day, they were aggregated to produce a single value for a given day (worst value for SOFA parameters and mean for others). Outliers were removed prior to aggregation by using predefined ranges for each measurement. When no measurements were available, we reported it as not available (NA). For patients on ECMO, we set $PaO_2/FiO_2$ ratio to NA; for patients on HD or CRRT, we set creatinine to NA, as these parameters are unreliable when patients are receiving these interventions. Full details are available in our code at https://github.com/NUSCRIPT/carpediem.

*Normalization.* To make measurements comparable between each other so that the clustering would discover the underlying structure of the data, we performed feature normalization. We used percentile normalization for all features and set tied values to the average percentiles.

*Correlation weighting.* Some measurements in our dataset displayed a high correlation due to mathematical or physiological coupling (e.g., plateau pressure, PEEP and lung compliance; PaCO2 and bicarbonate). To make such correlated measurements contribute comparable value to the difference between data points as an independent measurement, we applied correlation weighting (Supplemental Figure 2B). We defined related measurement groups as measurements that have Pearson correlation greater than 0.7 with at least one other measurement. We combined related features to a single feature by taking the mean percentile across the related features and then considering the percentile across these new values.

*Distance function.* We created a similarity matrix between all pairs of patient-days by computing the Pearson correlation among features that had values present for both patient-days after correlation weighting (Supplemental Figure 2C). We computed Euclidean distances on the similarity matrix.

*Clustering and the number of clusters.* We were interested in describing the highest number of clinical states that were clinically interpretable and produced reasonable between-cluster separation in mortality rates. After computing the distance metric between all pairs of data points, we performed hierarchical clustering using Ward's method[5] to acquire a dendrogram. To estimate what number of clusters best describes our data, we assessed how data points were grouped into 3 up to 40 clusters using a custom metric for differential mortality and by visually inspecting progressive splits of the dendrogram on a heatmap. We posited that clinically relevant clustering of patient-days would reveal clinical states that are associated with differential mortality. Thus, our custom metric was generated by computing the fraction of all pairs of clusters that had statistically significant differences in mortality using Fisher's exact test with $p<0.01$. Cluster mortality was computed as the number of patients with 'Died' binarized outcome among all patients who had at least one patient-day assigned to a cluster.

*Alternative clustering strategies.* We called the above data processing and clustering strategy "Similarity." We also tested two additional strategies, which we called "Ranked-Euclidean" and "Normalized-Euclidean." The "Ranked-Euclidean" strategy differs from "Similarity" by performing correlation weighting differently and by using a different distance function. "Normalized-Euclidean" strategy had alternative feature normalization, correlation weighting, and distance function procedures. We used the same methods as described in *Clustering and the number of clusters* to find the best clustering strategy, and also to choose the optimal threshold for highly correlated features.

*“Ranked-Euclidean” alternative strategy.* Highly correlated features were reweighted by dividing feature values by square root of the features’ group sizes. We computed distances between patient-days as Euclidean distance directly on the normalized and reweighted features, without computing a similarity matrix. We used an NA-robust implementation of Euclidean distances as described by Dixon,[6] available in *scikit-learn*.[7]

*“Normalized-Euclidean” alternative strategy.* For flag and score features, we performed percentile normalization as above. For other features, we applied additional steps. We applied winsorizing with a limit of 0.01, either one-sided or two-sided, depending on the measurement boundaries (e.g., oxygen saturation has a natural maximum of 100%, so we adjusted only the lowest 1% of values.). For features that displayed exponential distribution, we performed $\log_2$ transformation. We then linearly normalized values to a 0–1 range. We reweighted correlated features and computed distances between all patient-days as in the “Ranked-Euclidean” strategy.

*Robustness against exclusion of individual patient hospitalizations*. To evaluate the robustness of our clustering to minor perturbations, we randomly removed 100 different hospitalizations and performed the “Similarity” strategy. Across these randomizations, we found that individual patient-days would fall into clusters with similar associated mortality (Supplemental Figure 4B).

*Applicability of distances as metric for clustering.* NA-robust implementation of Euclidean distances used for the “Ranked-Euclidean” and “Normalized-Euclidean” does not guarantee preservation of the triangle inequality. To test the behavior of these strategies, we randomly sampled 10 million triplets of patient-days. We observed 0, 262, and 447 triplets that violated the triangle inequality when following the “Similarity,” “Ranked-Euclidean,” and “Normalized-Euclidean” strategies, respectively.

*Visualization*. To visualize clusters as distinct and recognizable clinical states, we ordered the clusters by increasing mortality, summarized clinical measurements for each cluster for each group of clinical measurements (neurologic, respiratory, shock, renal, inflammatory, and ventilator instability) and plotted the result as a heatmap (Figure 2B). Each measurement contributed to its group either directly (higher-is-worse) or inversely (lower-is-worse) so that all groups have higher-is-worse semantics. The four flags (ECMO, intubation, CRRT, and HD) were weighted with their original 0/1 score, whereas other parameters were weighted with their post-normalization value.

To visualize patient-days as clusters, we used the UMAP algorithm[8] to obtain 2D representation of our 44-dimensional dataset (Figure 2C). To overcome UMAP's inability to handle NAs, we disabled the *check_array* function, computed the kNN graph based on our predefined distance matrix (see *Distance function* above), and obtained UMAP 2D embedding by using the precomputed kNN graph. For final visualization we used 7 nearest neighbors and UMAP hyperparameters min_dist=0.2.

All plots were created in Python with matplotlib version 3.4.3 and seaborn version 0.11.2 libraries. Full details are available in our code at https://github.com/NUSCRIPT/carpediem.

*Data browser and example trajectory generation.* Trajectories formed by transitions between *CarpeDiem*-defined clinical states are a valuable resource for clinicians. To facilitate data exploration, reflection, and insight, we developed an interactive visualization for our dataset. Currently, the interface allows for patient-based data exploration; the user can select a patient and view the clinical states associated with each day of the patient's ICU stays, together with the clinical measurements. BAL sample and pneumonia episode information (see below) is overlaid onto the patient trajectory timeline. We used d3.js version 7.1.1 to create the interactive visualization. One of the patient trajectories is plotted in Figure 2D with relevant clinical events manually annotated. A demonstration browser can be viewed at https://nupulmonary.org/carpediem/, and a full browser will be available on PhysioNet.

*Modeling.* We used XGBoost[9] to model outcomes based on clinical features taking the worst values from the first two days in the patient's stay, consistent with commonly used ICU prediction scores. To this baseline model, we added a flag noting whether patients developed VAP during hospitalization and a flag noting whether this VAP's outcome was indeterminate or not cured (as opposed to cured). Confidence intervals were generated via bootstrapping. Models were trained using an 80/20 train/test split.

*Description of clinical adjudication process*. Outcomes for bacterial VAP episodes were adjudicated at day 7-8, day 10, and day 14 following the diagnostic BAL procedure. In total, 9,850 patient-days occurred following the SCRIPT enrollment. Cure was defined as the ability to survive beyond the duration of antibiotic treatment, the ability to remain off of antibiotics for 48 hours without recurrence or superinfection pneumonia, disappearance of the causative pathogen from BAL fluid or the absence of subsequent samples, absence of bacterial complications (e.g., empyema, lung abscess, endocarditis), and improvement in the clinical manifestations of pneumonia; successful extubation or ventilator liberation was considered as a cure. Failure was considered if the patient died during the antibiotic treatment course, the development of pneumonia led to a shift to comfort-only care, the patient developed complications such as empyema, lung abscess, or endocarditis or a persistent need for vasopressors or hemodynamic instability until a change in antibiotics. Specific failure modes included persistence, defined as interval recovery of the causative pathogen in a respiratory tract specimen, blood, or pleural fluid or development of an abscess/cavity, empyema, or endocarditis. Failure due to superinfection was defined as recovery of a new pneumonia pathogen while being treated for pneumonia. Indeterminate status indicated persistent inflammation (BAL fluid neutrophilia, fever, elevated white blood cell count without other explanation) or respiratory failure without demonstration of persistence or superinfection as defined above.

**Supplemental File 1. Standardized score sheet used by physician reviewers to adjudicate pneumonia episodes.**

**Separate file.**

**Supplemental File 2. The NU SCRIPT Study Investigators.**

**Separate file.**

| | Overall | Non-Pneumonia Control | Other Pneumonia | Other Viral Pneumonia | COVID-19 |
|---|---|---|---|---|---|
| **n** | 585 | 93 | 252 | 50 | 190 |
| **Age, median [Q1,Q3]** | 62.0 [51.0,72.0] | 60.0 [49.0,70.0] | 65.0 [52.0,73.0] | 59.5 [52.2,69.8] | 61.0 [51.0,70.0] |
| **Ethnicity, n (%)** | | | | | |
| **Hispanic or Latino** | 113 (19.3) | 11 (11.8) | 22 (8.7) | 11 (22.0) | 69 (36.3) |
| **Not Hispanic or Latino** | 422 (72.1) | 75 (80.6) | 211 (83.7) | 35 (70.0) | 101 (53.2) |
| **Unknown or Not Reported** | 50 (8.5) | 7 (7.5) | 19 (7.5) | 4 (8.0) | 20 (10.5) |
| **Gender, n (%)** | | | | | |
| **Female** | 239 (40.9) | 46 (49.5) | 100 (39.7) | 24 (48.0) | 69 (36.3) |
| **Male** | 346 (59.1) | 47 (50.5) | 152 (60.3) | 26 (52.0) | 121 (63.7) |
| **Race, n (%)** | | | | | |
| **Asian** | 17 (2.9) | 6 (6.5) | 7 (2.8) | 2 (4.0) | 2 (1.1) |
| **Black/African American** | 116 (19.8) | 17 (18.3) | 54 (21.4) | 8 (16.0) | 37 (19.5) |
| **Unknown or Not Reported** | 111 (19.0) | 14 (15.1) | 32 (12.7) | 6 (12.0) | 59 (31.1) |
| **White** | 341 (58.3) | 56 (60.2) | 159 (63.1) | 34 (68.0) | 92 (48.4) |
| **Smoking status, n (%)** | | | | | |
| **Current Smoker** | 48 (8.2) | 9 (9.7) | 30 (11.9) | 5 (10.0) | 4 (2.1) |
| **Never Smoker** | 230 (39.3) | 40 (43.0) | 94 (37.3) | 22 (44.0) | 74 (38.9) |
| **Past Smoker** | 150 (25.6) | 25 (26.9) | 78 (31.0) | 19 (38.0) | 28 (14.7) |
| **Unknown Smoking Status** | 157 (26.8) | 19 (20.4) | 50 (19.8) | 4 (8.0) | 84 (44.2) |
| **BMI, median [Q1,Q3] *** | 28.0 [24.0,33.2] | 27.1 [24.0,32.6] | 26.6 [22.3,32.4] | 26.7 [23.8,31.3] | 30.6 [26.6,35.8] |
| **Admit APS score, median [Q1,Q3]** | 89.0 [64.0,107.0] | 90.0 [62.0,105.0] | 88.0 [66.0,109.0] | 86.0 [64.2,100.0] | 90.0 [61.2,106.8] |
| **Admit SOFA score, median [Q1,Q3]** | 11.0 [8.0,13.0] | 11.0 [8.0,14.0] | 11.0 [8.0,14.0] | 10.0 [7.0,13.0] | 11.0 [8.2,13.0] |
| **Cumulative ICU days, median [Q1,Q3] **** | 14.0 [6.0,26.0] | 8.0 [4.0,17.0] | 10.0 [5.8,20.0] | 11.0 [7.5,19.8] | 24.0 [14.0,36.8] |
| **Number of ICU stays, median [Q1,Q3]** | 1.0 [1.0,1.0] | 1.0 [1.0,1.0] | 1.0 [1.0,2.0] | 1.0 [1.0,2.0] | 1.0 [1.0,1.0] |
| **No tracheostomy, n(%)** | 434 (74.2) | 78 (83.9) | 204 (81.0) | 43 (86.0) | 109 (57.4) |
| **Had tracheostomy, n(%)** | 151 (25.8) | 15 (16.1) | 48 (19.0) | 7 (14.0) | 81 (42.6) |
| **Cumulative intubation days, median [Q1,Q3] **** | 10.0 [4.0,23.0] | 5.0 [2.0,12.0] | 8.0 [4.0,18.0] | 9.0 [3.0,14.0] | 21.0 [10.0,35.0] |
| **Discharge disposition, n (%)** | | | | | |
| **Died ***** | 243 (41.5) | 37 (39.8) | 99 (39.3) | 20 (40.0) | 87 (45.8) |
| **Home** | 133 (22.7) | 27 (29.0) | 49 (19.4) | 10 (20.0) | 47 (24.7) |
| **LTACH** | 60 (10.3) | 6 (6.5) | 28 (11.1) | 4 (8.0) | 22 (11.6) |
| **Rehab** | 97 (16.6) | 11 (11.8) | 48 (19.0) | 12 (24.0) | 26 (13.7) |
| **SNF** | 33 (5.6) | 6 (6.5) | 18 (7.1) | 1 (2.0) | 8 (4.2) |
| **Hospice** | 19 (3.2) | 6 (6.5) | 10 (4.0) | 3 (6.0) | - |

**Supplemental Table 1. Demographics and outcomes data for the cohort, grouped by pneumonia category.**
* With the exception of one BMI value, all values were complete for every patient. Racial groups with fewer than five individuals were classified as ‘Unknown or Not Reported’ to protect patient anonymity.
**Total days intubated and total ICU days include only days at our hospital and do not capture intubation duration or ICU LOS at a transferring hospital.
*** Died included those who died or underwent lung transplantation for refractory respiratory failure.
BMI = body mass index, APS = Acute Physiology Score (score calculated from worst value within the first two ICU days), SOFA = Sequential Organ Failure Assessment (score calculated from worst value within the first two ICU days).

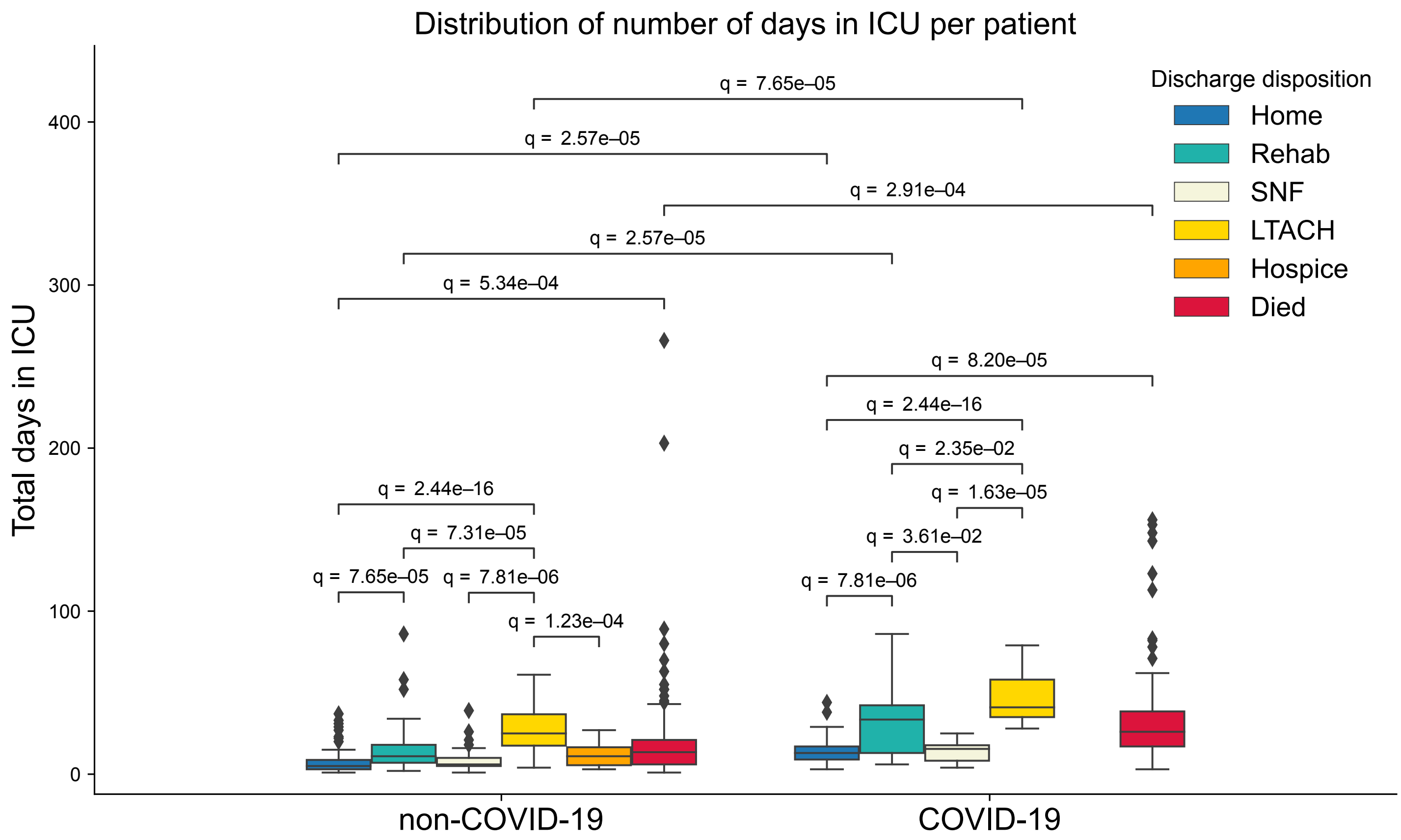


**Supplemental Figure 1. Length of stay by discharge disposition among patients with and without COVID-19.** Patients with severe SARS-CoV-2 pneumonia experienced significantly longer lengths of ICU stay for all discharge disposition groups, except Skilled Nursing Facility (SNF; not significantly different) and Hospice (no patients in SARS-CoV-2 group).

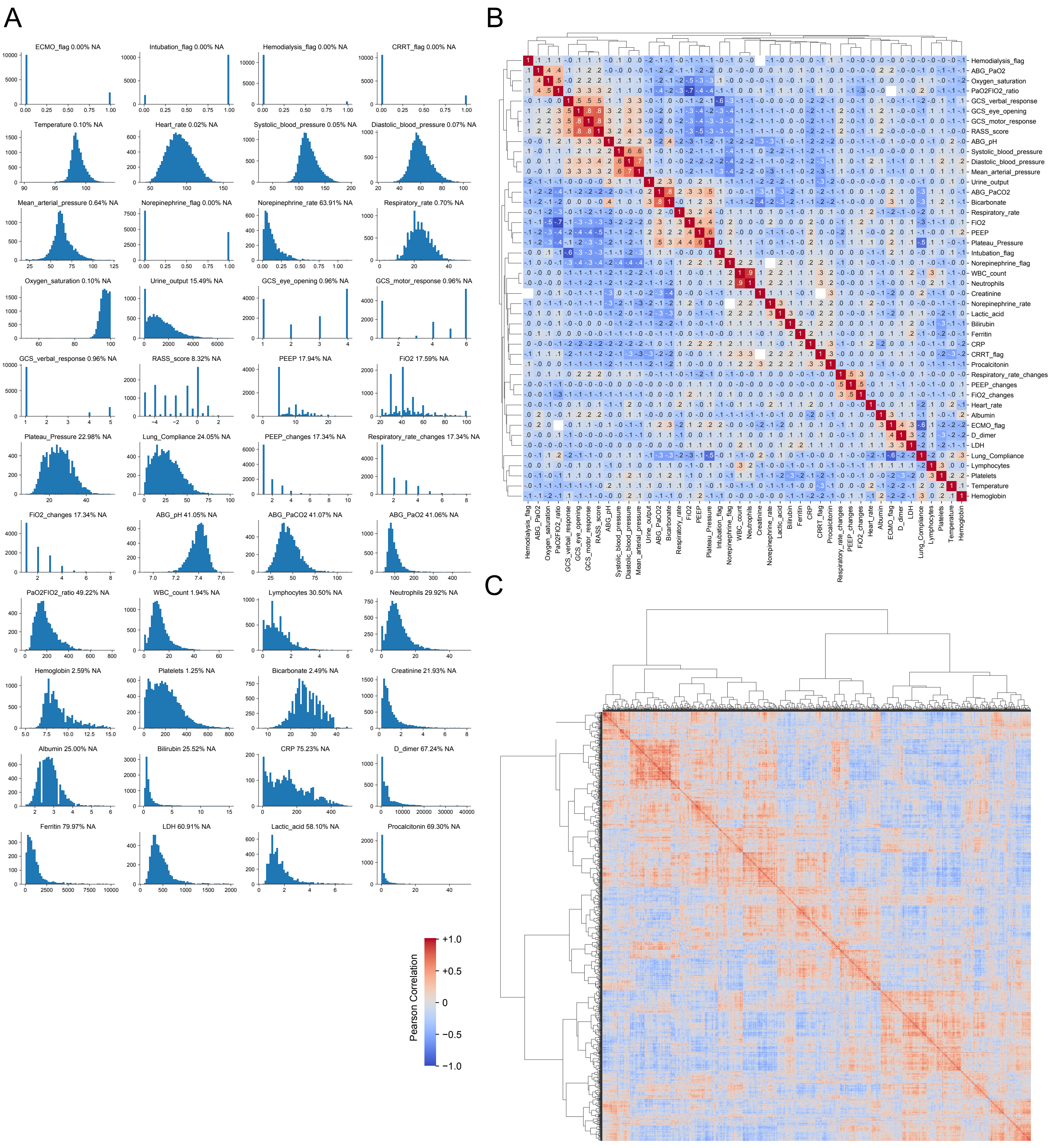


**Supplemental Figure 2. Clinical parameters and their correlation.** (**A**) Histogram display of 44 clinical values extracted from the EHR. (**B**) Pearson correlation matrix of clinical parameters; some measurements in our dataset displayed a high correlation due to mathematical or physiological coupling (e.g., plateau pressure, PEEP and lung compliance; PaCO2 and bicarbonate). (**C**) Pearson correlation between different patient-days as used as an intermediate step for the “Similarity” clustering strategy.

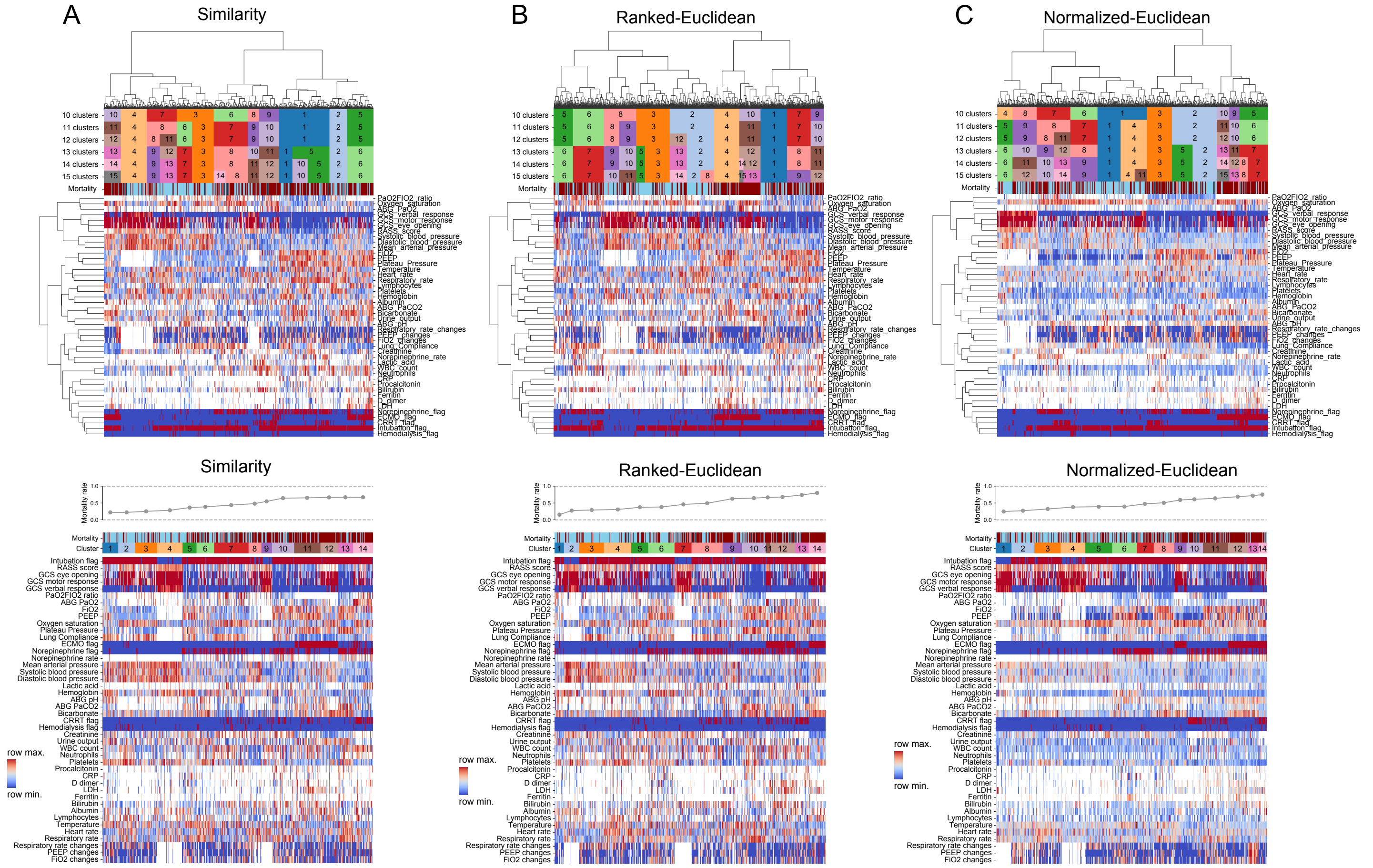

**Supplemental Figure 3. Comparing three different strategies for clustering.** (**A**) Similarity, (**B**) Ranked-Euclidean, and (**C**) Normalized-Euclidean strategies. For each method, hierarchical clustering of clinical parameters (rows) and columns (patient-days) is shown on the top, grouping patient-days into 10-15 separate clusters. The bottom panels show re-ordered clustering with columns organized into clusters and sorted by ascending cluster mortality and rows organized into physiologically similar groups. Cluster mortality is shown above the heatmaps.

A

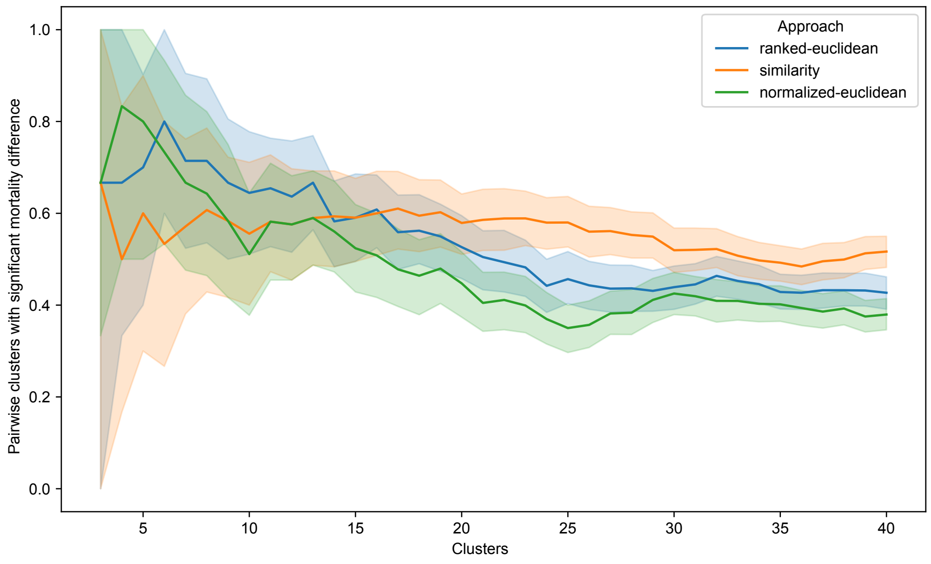


B

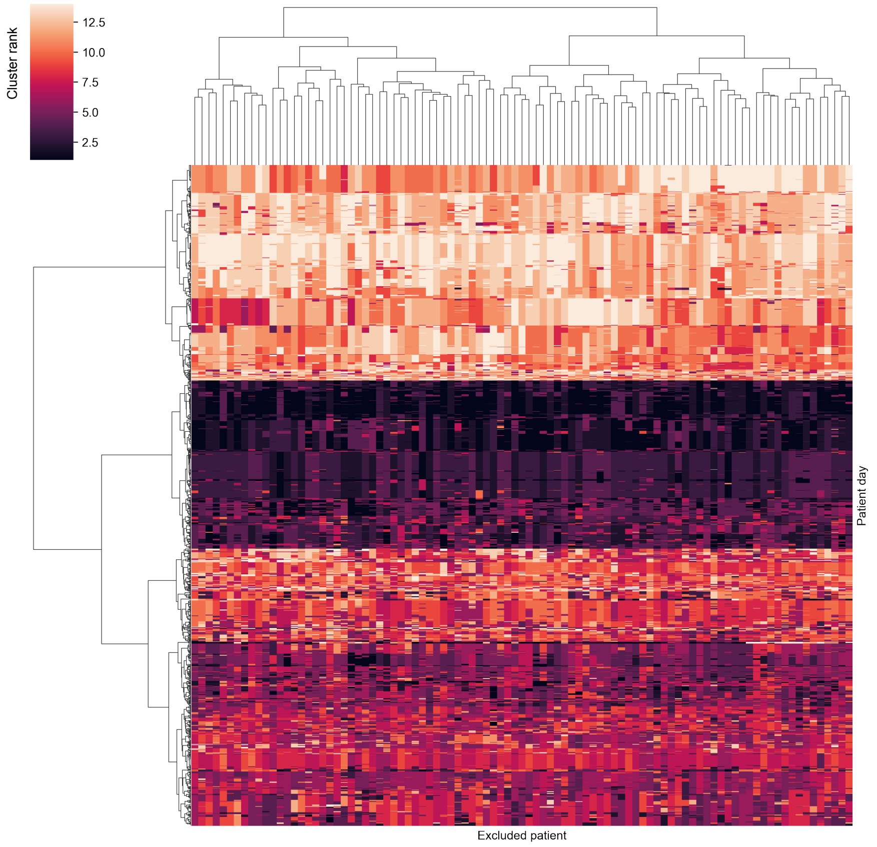


**Supplemental Figure 4. Cluster mortality differentiation and robustness against small data perturbations.** (**A**) Fraction of all possible pairs between two clusters that show a significantly different mortality at p<0.01. X-axis shows different cutoff for total number of clusters. Shaded area is bootstrapped 95 percentile. (**B**) Allocation of individual patient-days (rows) to clusters following 100 randomizations in which a single patient hospitalization has been excluded (columns). Cluster rank is the rank of cluster based on associated mortality with 1 being lowest and 14 being highest.

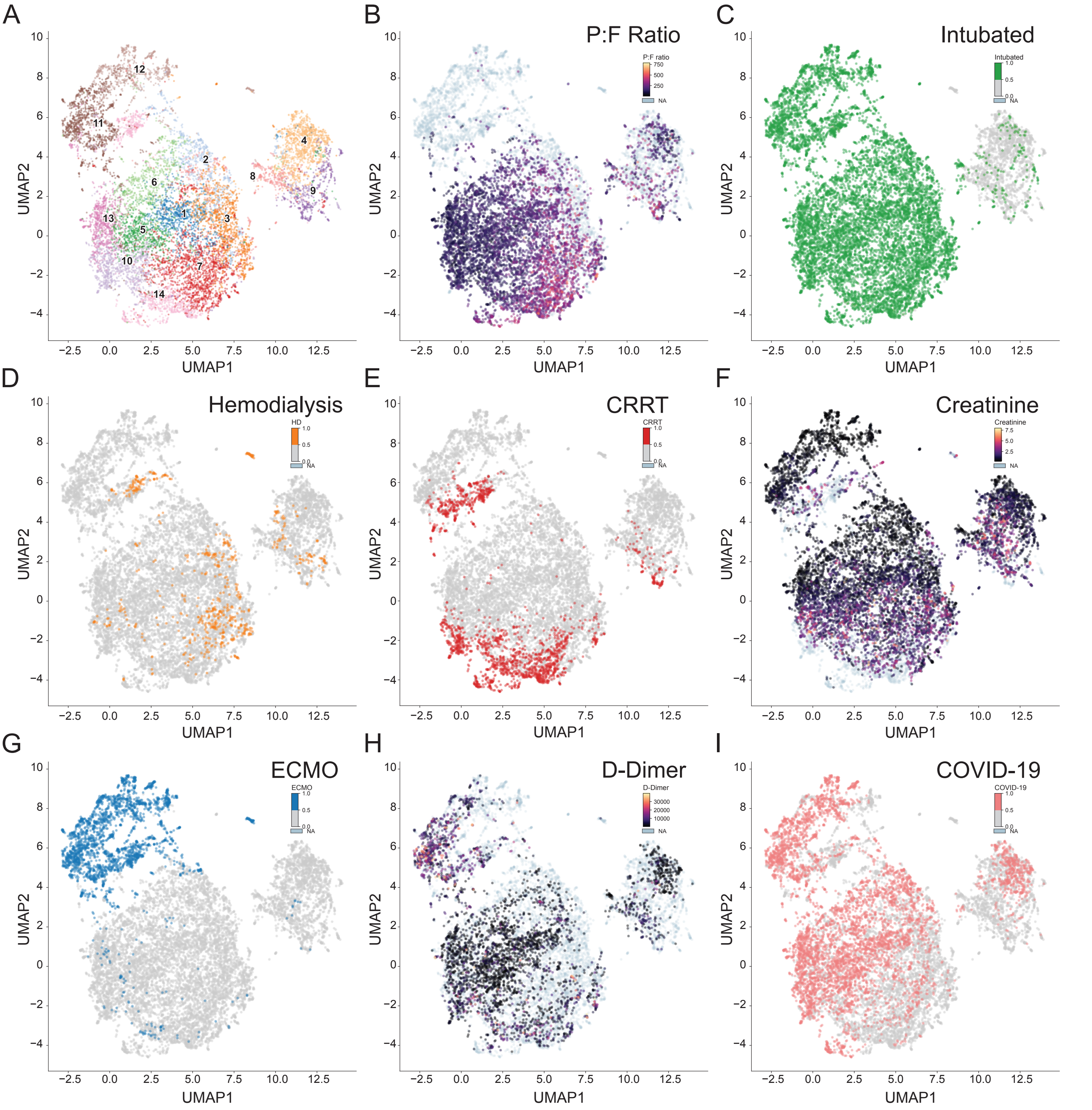


**Supplemental Figure 5. Feature plots (refers to Figure 2).** (**A**) Reference UMAP with numbered clusters. (**B-H**) Feature plots for individual parameters. (**I**) Patient-days from patients with COVID-19.

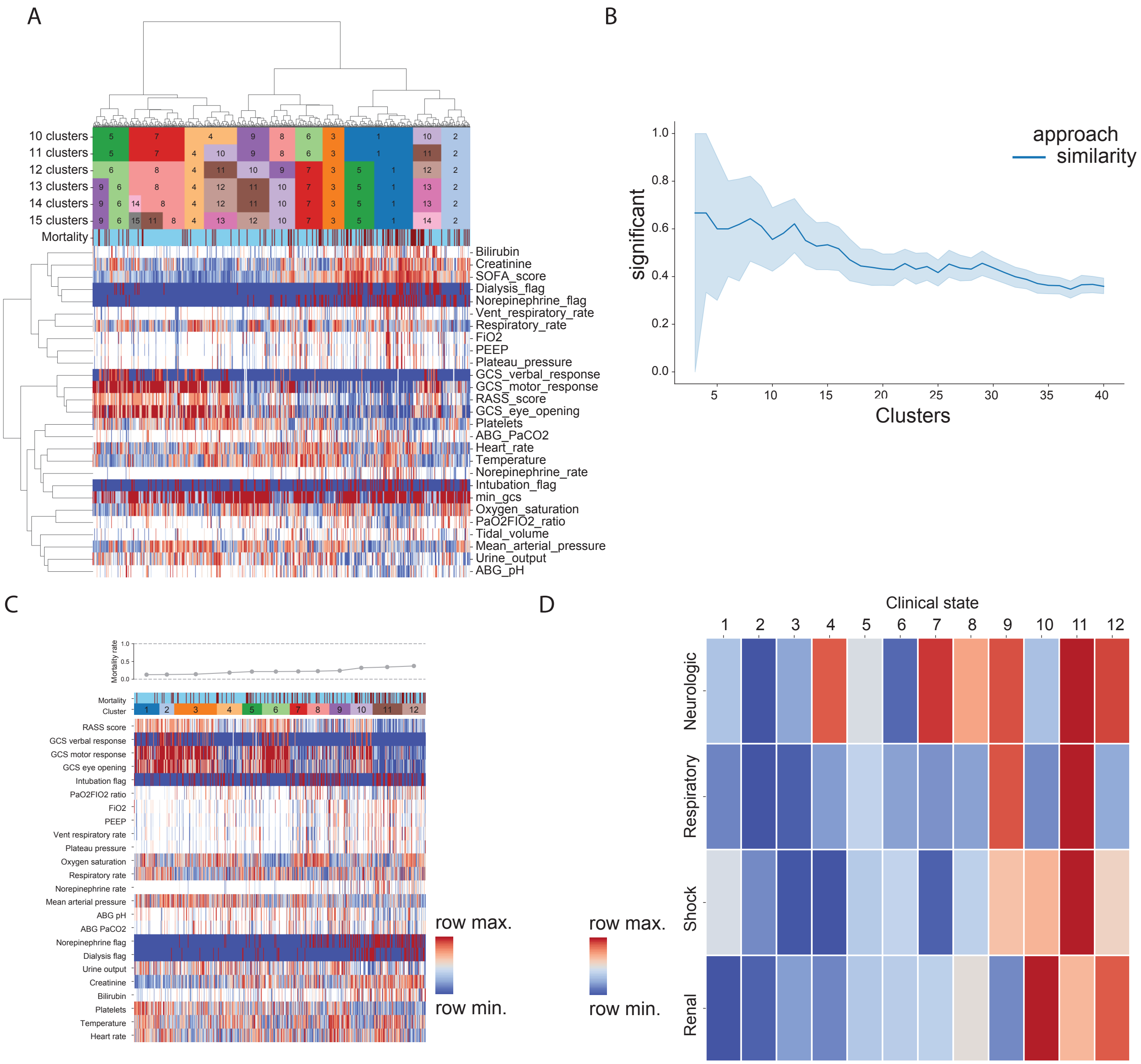


**Supplemental Figure 6. Application of *CarpeDiem* to the MIMIC-IV dataset.** (**A**) Hierarchical clustering of 27 clinical parameters (rows) with columns representing 15,642 ICU patient-days from 1,284 patients. (**B**) The fraction of pairs between clusters that show a significantly different mortality at different numbers of clusters. (**C**) Heatmap of data from (A) re-ordered from lowest to highest mortality, using 12 clusters. The top strip signifies hospital mortality of the patient shown in the column (blue = survived, red = died). The hospital mortality rate associated with each cluster is shown above the heatmap. (**D**) Heatmap of the composite signal from each cluster and physiologic group with ordering same as (C).

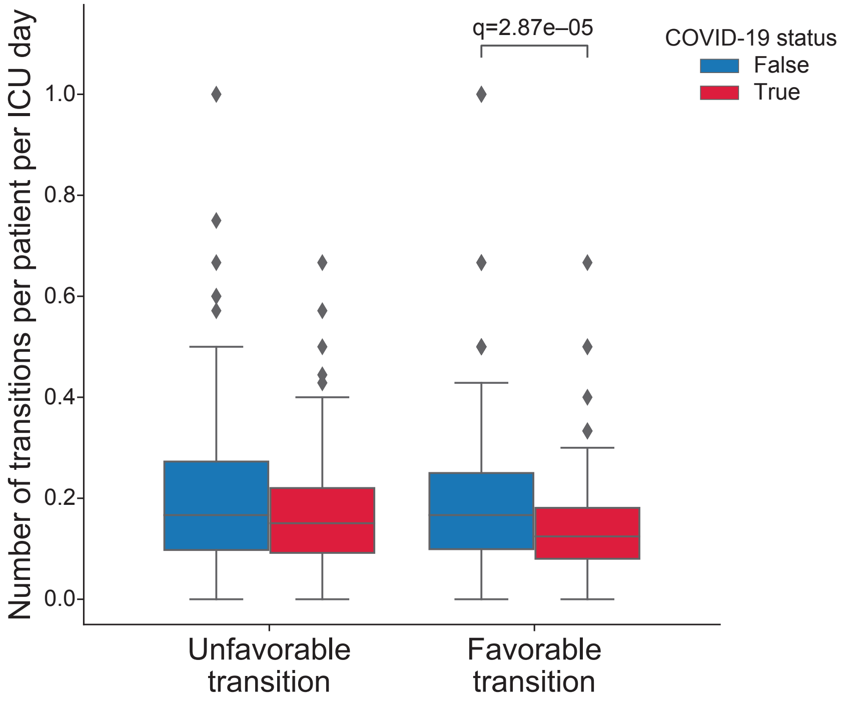


**Supplemental Figure 7. Favorability of transitions.** Normalized transitions by total ICU-days split by the favorability of transitions experienced by patients with or without COVID-19.

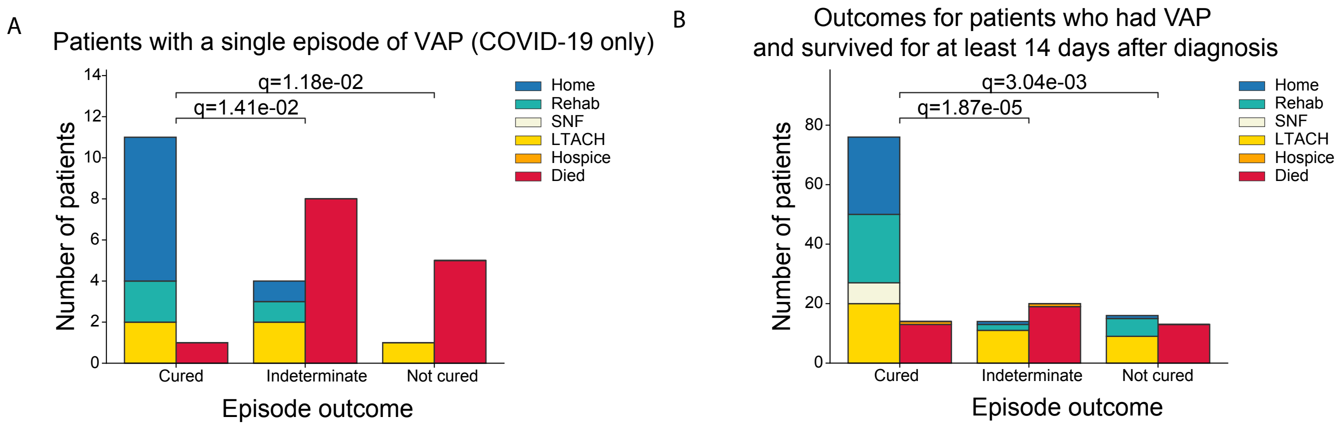


**Supplemental Figure 8. Unresolving VAP is associated with poorer outcomes.** (**A**) Mortality associated with a single episode of VAP among patients with COVID-19. (**B**) Outcomes for patients who did not die within 14 days of the start of their VAP episode. Outcomes are displayed in two columns: the first column aggregates favorable discharge dispositions (Home, Rehab, SNF, LTACH), the second column aggregates unfavorable discharges (Hospice, Died).

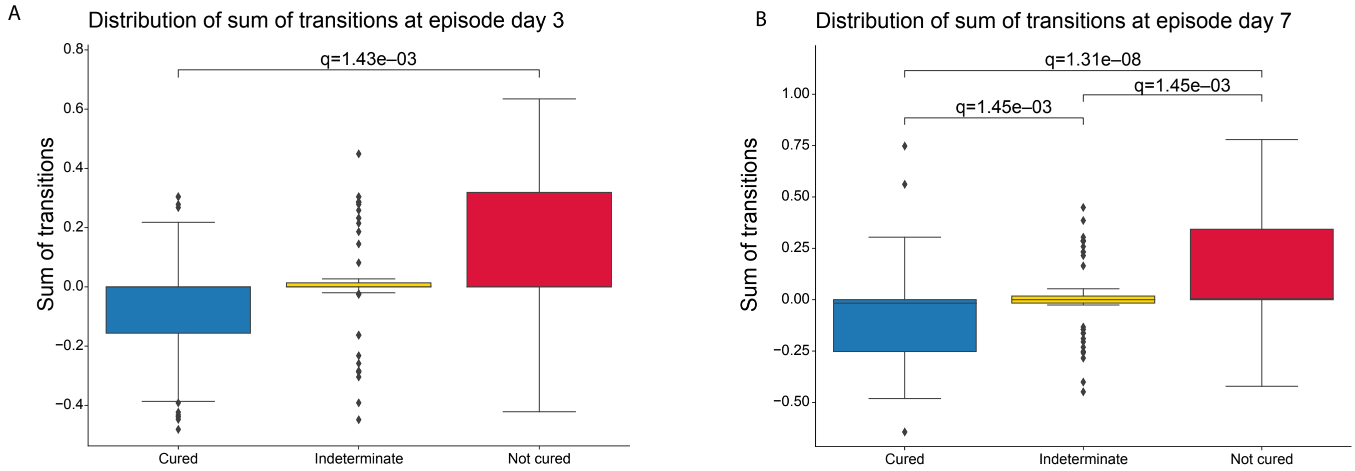


**Supplemental Figure 9. Trajectory analysis reveals statistically significant differences between cured, indeterminate, and not cured VAP episodes.** Summation of transitions between clinical states: negative values represent favorable transitions; positive values represent unfavorable transitions. (**A**) Significant differences between cured and not cured VAP episodes are revealed as early as day 3. (**B**) Day 7 has significant differences between all groups.

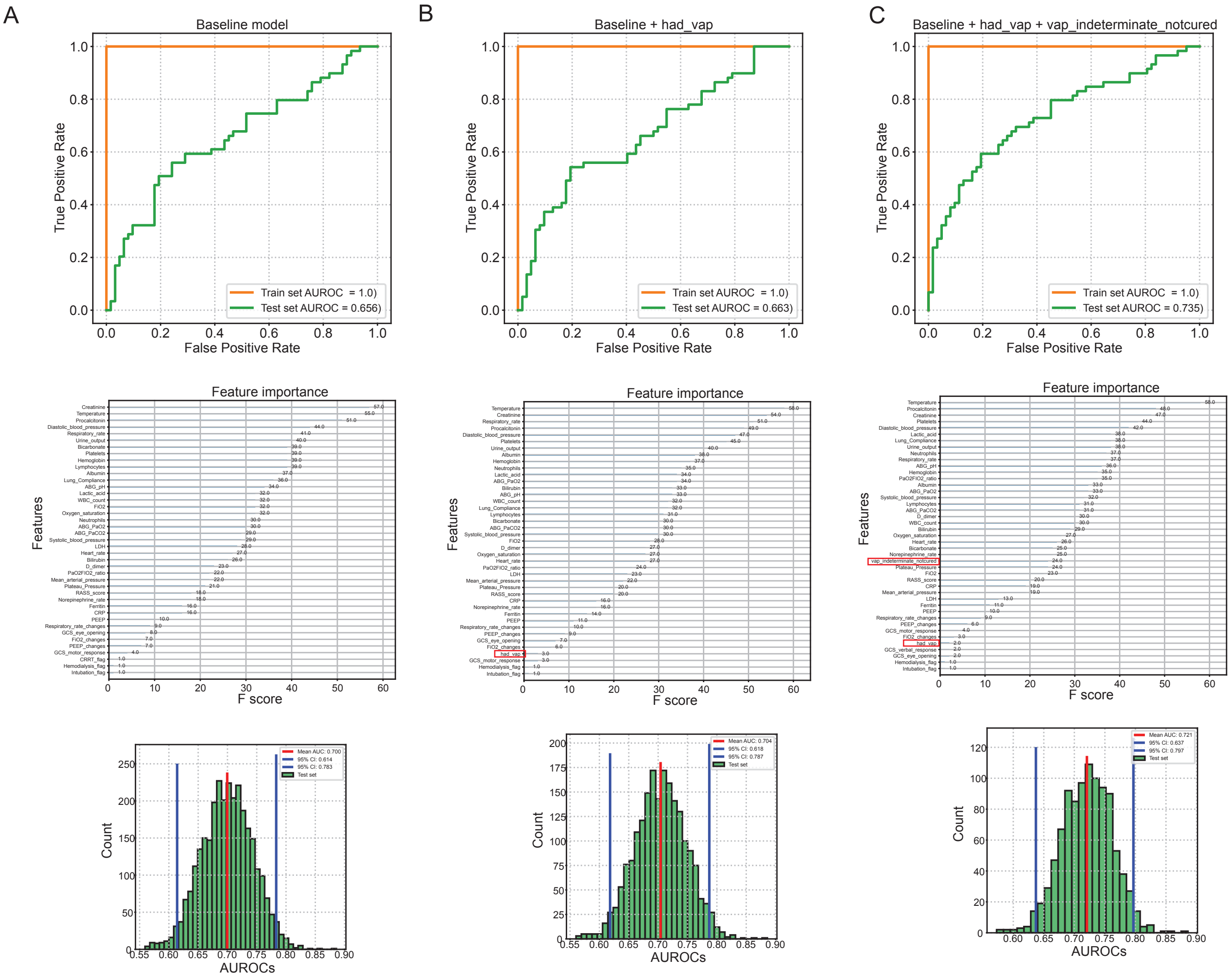


**Supplemental Figure 10. Gradient boosting analysis reveals minimal increase in predictive capability when VAP and VAP cure status are added to clinical parameters measured early in the ICU course.** (**A**) Area under the receiver operating characteristics (AUROC) curve values for clinical parameters (top) with corresponding feature importance (middle) in the gradient boosting analysis predicting hospital mortality. Bottom row shows confidence intervals of the AUROC curve obtained using bootstrapping. (**B**) Gradient boosting analysis adding a *had_vap* flag, to indicate a diagnosis of VAP during the ICU stay. (**C**) Gradient boosting analysis adding two flags, *had_vap* and *vap_interdeterminate_uncured*, to indicate a diagnosis of VAP during the ICU stay and a VAP outcome other than cured. Point estimates and confidence intervals in the analyses are (A) 0.656 [0.614,0.783], (B) 0.663 [0.618,0.787], and (C) 0.725 [0.637,0.797].

### Works Cited


1 Office for Civil Rights (OCR). Guidance regarding methods for DE-identification of protected health information in accordance with the Health Insurance Portability and Accountability Act (HIPAA) Privacy Rule. HHS.gov. 2012; published online Sept 7. https://www.hhs.gov/hipaa/for-professionals/privacy/special-topics/de-identification/index.html (accessed Sept 9, 2022).

2 Johnson A, Bulgarelli L, Pollard T, Horng S, Celi LA, Mark R. MIMIC-IV. 2022. DOI:10.13026/7VCR-E114.

3 Pollard TJ, Johnson AEW, Raffa JD, Mark RG. tableone: An open source Python package for producing summary statistics for research papers. *JAMIA Open* 2018; **1**: 26–31.

4 Starren JB, Winter AQ, Lloyd-Jones DM. Enabling a Learning Health System through a Unified Enterprise Data Warehouse: The Experience of the Northwestern University Clinical and Translational Sciences (NUCATS) Institute. *Clin Transl Sci* 2015; **8**: 269–71.

5 Ward JH. Hierarchical Grouping to Optimize an Objective Function. *J Am Stat Assoc* 1963; **58**: 236–44.

6 Dixon JK. Pattern Recognition with Partly Missing Data. *IEEE Trans Syst Man Cybern* 1979; **9**: 617–21.

7 Pedregosa F, Varoquaux G, Gramfort A, *et al.* Scikit-learn: Machine Learning in Python. *J Mach Learn Res* 2011; **12**: 2825–30.

8 McInnes L, Healy J, Melville J. UMAP: Uniform Manifold Approximation and Projection for Dimension Reduction. arXiv [stat.ML]. 2018; published online Feb 9. http://arxiv.org/abs/1802.03426.

9 Chen T, Guestrin C. XGBoost: A Scalable Tree Boosting System. In: Proceedings of the 22nd ACM SIGKDD International Conference on Knowledge Discovery and Data Mining. New York, NY, USA: Association for Computing Machinery, 2016: 785–94.
